# Antibody response and intra-host viral evolution after plasma therapy in COVID-19 patients pre-exposed or not to B-cell depleting agents

**DOI:** 10.1101/2022.04.24.22274200

**Authors:** David Gachoud, Trestan Pillonel, Tsilimidos Gerasimos, Dunia Battola, Dominique Dumas, Onya Opota, Stefano Fontana, Peter Vollenweider, Oriol Manuel, Gilbert Greub, Claire Bertelli, Nathalie Rufer

## Abstract

**Background:** Administration of plasma therapy may contribute to viral control and survival of COVID-19 patients receiving B-cell depleting agents that hinder the endogenous humoral response. However, little is known on the impact of anti-CD20 pre-exposition and the use of different sources of plasma (convalescent versus vaccinated) on the kinetics of SARS-CoV-2-specific antibodies and viral evolution after plasma therapy.

**Methods:** Eligible COVID-19 patients (n = 36), half of them after anti-CD20 targeted therapy, were treated with therapeutic plasma from convalescent (n = 17) or mRNA-vaccinated (n = 19) donors. Each plasma-transfused patient was thoroughly monitored over time by anti-S IgG quantification and whole-genome SARS-CoV-2 sequencing.

**Results:** The majority of anti-CD20 pre-exposed patients (15/18) showed progressive declines of anti-S protein IgG titers following plasma therapy, indicating that they mostly relied on the passive transfer of anti-SARS-CoV-2 antibodies. Such antibody kinetics correlated with prolonged infection before virus clearance, contrasting with the endogenous humoral response predominantly present in patients who had not received B-cell depleting agents (15/18). No relevant differences were observed between patients treated with plasma from convalescent and/or vaccinated donors. Finally, 4/30 genotyped patients showed increased intra-host viral evolution and 3/30 included 1 to 4 spike mutations, potentially associated to immune escape.

**Conclusions:** Convalescent and/or vaccinated plasma therapy may provide anti-SARS-CoV-2 antibodies and clinical benefit to B-cell depleted COVID-19 patients. Only a limited number of patients acquired viral mutations prior to clinical recovery, yet our study further emphasizes the need for long-term surveillance for intra-host variant evolution, to guide best therapeutic strategies.

## INTRODUCTION

Coronavirus disease 2019 (COVID-19) disproportionally affects immunocompromised patients, in the context of their underlying disease, high prevalence of comorbidities, and/or related treatment (1). Hematological malignancies and solid tumors have been consistently associated with increased risk of COVID-19 complications and death (2-5). Repeated administration of anti-CD20 monoclonal antibody (e.g. rituximab), an effective treatment for B-cell cancers or inflammatory autoimmune diseases, which leads to B-cell lymphopenia and hypogammaglobulinemia, is also marked by a more severe COVID-19 course (6-8) and impaired anti-SARS-CoV-2 antibody response, elicited by infection or vaccination (9, 10).

Neutralizing antibodies represent an important correlate of recovery following SARS-CoV-2 infection (11). Consequently, convalescent plasma therapy (CP), obtained from donors who have recovered from COVID-19 and containing anti-SARS-CoV-2 neutralizing antibodies, has been under massive investigation as reported in large randomized controlled trials (12-18). In immunocompetent COVID-19 patients with high-risk factors for severe disease progression, treatment with CP has shown clinical benefit when given early (within 72 hours after the onset of symptoms) and with high titers of neutralizing antibodies (12, 13). Similar observations were made when REGN-COV2, a neutralizing monoclonal antibody (mAb) cocktail was administrated early in the disease course and in seronegative individuals (19). Most of these trials, however, failed to demonstrate a therapeutic benefit of CP, once COVID-19 patients were hospitalized with an already-established severe pneumonia (14-18).

The usefulness of plasma therapy is more substantial in immunodeficient patients. There is growing evidence from cohort studies and case-series, that CP therapy in frail immunosuppressed individuals, unable to mount effective anti-SARS-CoV-2 antibody responses, reduces viral load and improves clinical symptoms, even when given late after initial diagnosis (20-25). Accordingly, these findings suggest that the administration of plasma with high neutralizing antibody titers is a safe and effective treatment for immunosuppressed patients (3, 26, 27).

Patients with immunosuppression are also at specific risk for a protracted infection with SARS-CoV-2 (28). In an initial report, Aydillo *et al*. showed no major changes in the consensus sequences of the original virus strain from serial sample genomes of 11 immunosuppressed patients, including patients treated with CP (29). However, accumulation of SARS-CoV-2 mutations have been documented in sporadic case-reports of long-term infected immunocompromised hosts (30-34). While this phenomenon does not seem to be very common (26, 35), prolonged viral replication in the context of an inadequate immune response may facilitate the emergence of divergent escape variants (28).

A key issue of CP therapy relates to the wide heterogeneity of neutralizing antibody titers found within CP units from recovered individuals (36). The rapid decay of circulating antibody titers within 2 to 3 months after viral infection (37) strongly limits the window of opportunity to collect high-dose anti-SARS-CoV-2 IgG titers from convalescent donor plasma (38). For instance, the supply of CP from one center revealed that high titer collections, as defined by the US Food and drug Administration (FDA), accounted for only about 20% of plasma donations (39). In turn, SARS-CoV-2 antibody responses induced after the second dose of mRNA vaccines are found to be similar to or even higher than the average values from convalescent serum samples (40, 41). Moreover, planning plasmapheresis from individuals who have scheduled their vaccination date is logistically easier than from COVID-19 recovered donors. Assuming that the main criteria of plasma efficacy is to provide the highest antiviral antibody titers, this argument supports the use of plasma from non-COVID-19 healthy adults who had recently received the second dose of an mRNA-based vaccine.

Here, we describe the long-term outcomes of 36 patients with acquired immunodeficiencies (32/36; 89%) or high-risk factors (4/36; 11%) after treatment with convalescent plasma (CP, n = 17) or vaccinated plasma (VP, n = 19), between November 2020 and July 2021. Half of the patients (18/36) had received or were still under anti-CD20 therapy (e.g. rituximab, obinutuzumab). The aims of this study were to determine (i) the feasibility of using different sources of plasma (CP versus VP), (ii) the impact of anti-CD20 pre-exposition on anti-SARS-CoV-2-specific antibody kinetics and (iii) the rate of viral evolution and immune escape after plasma therapy in immunosuppressed patients. Therefore, each patient was thoroughly monitored over time by anti-S IgG quantification and whole-genome SARS-CoV-2 sequencing.

## ETHICS STATEMENT

The study protocol was reviewed and approved by the Medical Board Committee of the Lausanne University Hospital (Lausanne, Switzerland) and the Cantonal Ethics Committee for Clinical Research (CER-VD; Lausanne, Switzerland). Each patient or a legally authorized representative provided informed consent prior to plasma transfusion and for the retrospective data collection. Each donor gave written consent for plasma donation and collection of sera, according to regulations of the Swiss Blood Transfusion Services.

## METHODOLOGY

### Study design

All patients described in this case series were treated with either CP (n = 17) or VP (n = 19) between November 27, 2020 and July 28, 2021 under an experimental therapy protocol available as compassionate use only, according to the Swiss Federal Law on Therapeutic Products (LPTh). Eligible patients (>18 years of age) had a PCR-confirmed diagnosis of SARS-CoV-2 infection, a documented onco-hematological diagnostic (n = 22) or an autoimmune disease (n = 5) and/or a solid organ transplant (n = 5) and/or active solid tumor malignancy (n = 3) and were hospitalized with mild to severe COVID-19 according to the WHO classification. We included 4 additional non-immunocompromised patients with high-risk factors for severe COVID-19 and nosocomial infection < 72h post symptoms or diagnosis (12). Patients included in this analysis were alive on day 7 after plasma transfusion.

### Plasma collection and preparation

Convalescent plasma was collected from 32 male donors (CP), who had fully recovered for at least 28 days after COVID-19 onset and presented an anti-S protein-specific IgG response, ranging from 29.4-135.6 ratio, with an average ratio of 79 using an in-house developed Luminex assay (42), correlating to neutralizing titers (43, 44). Due to the difficulties to obtain high-titer plasma from convalescent donors, we collected from March, 1^st^, 2021 onwards, plasma from 24 non-COVID-19 male donors (VP), who had received their second dose of an mRNA-based vaccine (Moderna or Pfizer) and exhibited an anti-S protein IgG response ranging from 73.3-143.7.6 ratio, with an average ratio of 118 by Luminex. Since June 2021, we harvested plasma from convalescent male donors, boosted with an mRNA-based vaccine (Moderna or Pfizer) after COVID-19 infection (CP/VP). In addition, anti-S IgG titers was assessed in each donation by ELISA from EuroImmun (CP, [ranging from 1.34-10 S/CO, average ratio of 5] ; VP and CP/VP >8 S/CO). All plasma donors (18-65 years old) were eligible for blood donation according to the requirements of the Blood Transfusion Services, Swiss Red Cross. After apheresis collection, the leukocyte-depleted plasma was treated for pathogen-inactivation (Intercept blood system, Cerus Corporation) and standard testing according to the current regulations in Switzerland (Blood Transfusion Services CRS and Swiss Federal Act on Medical Products). The plasma was further separated into three units (200+/-20 ml each) within 24 hours and kept frozen at -25°C. Plasma recipients were transfused either with four units of ABO-compatible CP (from ≥2 different donors), given on two consecutive days or with two units of ABO-compatible VP or CP/VP (from 2 different donors whenever possible), given on the same day. Each unit was administrated over a 45-minute period, without any adverse event.

### Virus detection by qRT-PCR

The SARS-CoV-2 RNA was detected in various clinical specimens by real-time PCR using the different platforms available in our diagnostic laboratory, namely a fully automated molecular diagnostic platform MDx platform, the Xpert platform (Cepheid, Ca, USA), the cobas 6800 platform and the cobas Liat platform (Roche Basel, CH), as described in (45, 46). All obtained Ct values were converted to viral loads based on plasmids positive controls, as reported in (46).

### Anti-S-protein specific IgG titers by Luminex assay

Sera from individuals at the time of plasma donations (CP versus VP) and sera from patients at different time-points after plasma transfusion were collected and characterized for anti-spike protein (S1) IgG titers using an in-house developed Luminex assay and performed as previously described (42). MFI signal of each donor serum or plasma patient sample was divided by the mean signal for the negative control samples yielding a MFI ratio (42).

### SARS-CoV-2 whole genome sequencing

RNA from clinical samples (nasopharyngeal or mouth swabs) were extracted using MagnaPure (Roche, Switzerland) and processed with the CleanPlex SARS-CoV-2 panel as previously described (47). Briefly, the CleanPlex SARS-CoV-2 protocol generates 343 amplicons ranging from 116 to 196 bp (median, 149 bp), distributed into two pools. All samples were sequenced using 150-bp paired-end protocol on a MiSeq (Illumina, USA). Reads were processed using GENCOV (https://github.com/metagenlab/GENCOV), a modified version of CoVpipe (https://gitlab.com/RKIBioinformaticsPipelines/ncov_minipipe). Variant calling was performed with Freebayes (parameters: –min-alternate-fraction 0.1 –min-coverage 10 –min-alternate-count 9) (48). Positions covered by less than 10 reads were set to N (unknown) if they were not identified as part of a short deletion by Freebayes. Only variants supported by at least 70% of mapped reads were considered to build consensus genomes. The consensus sequence was generated with bcftools (49), was checked using our in-house quality control (50) and assigned to SARS-CoV-2 lineages with pangolin (51).

### Intra-host mutation rate and phylogenetic analyses

Single nucleotide variants and indels supported by more than 10% of the reads were compared between sequenced SARS-CoV-2 genomes of each patient. The mutation rate was calculated as the total number of variants supported by >10% of reads present in one or multiple sequenced genomes and absent from the other sequenced genome(s). In addition, the rate of mutations reaching fixation was calculated as the total number of variants supported by >=70% of the reads in the last sequenced sample and absent (or supported by <70% of the reads) from the first sequenced genome divided by the time interval (in days) between the first and the last sample. Phylogenies were built using Nextstrain and the ncov workflow (https://github.com/nextstrain/ncov, (52)) including publicly available genomes sequenced in Switzerland (GISAID database version 2022-01-21; https://www.eurosurveillance.org/content/10.2807/1560-7917.ES.2017.22.13.30494?crawler=true). Sequencing reads were submitted to the International Nucleotide Sequence Database Collaboration (INSDC), whereas consensus genome sequences were submitted to GISAID and onto the Swiss Pathogen Surveillance Plateform.

## RESULTS

### Characteristics of patients treated with convalescent or vaccinated plasma

Seventeen patients (6 female/11 male) with acquired immunodeficiencies due to hematological malignancy (88%) or autoimmune disease (12%) were treated with convalescent plasma (CP) from November 27, 2020 to March 17, 2021 (**Supp. Table 1**). Among them, 12 (71%) had received or still were under anti-CD20 therapy. As the Swiss vaccination campaign started in early 2021, it became possible to collect plasma from SARS-CoV-2 vaccinated regular blood donors without a previous history of SARS-CoV-2 infection. Consequently, vaccinated plasma (VP) was administrated to 19 patients (9 female/10 male) with hematological cancer (37%) or non-hematological disease (42%; autoimmune disease, organ transplant or solid tumor) or with high-risk factors for severe COVID-19 progression (21%), from March 09, 2021 to June 22, 2021 (**Supp. Table 1**). Patients treated with VP were predominantly infected by the alpha variant B.1.1.7, the variant of concern spreading rapidly in Switzerland at that time (from January to June 2021), in contrast to CP treated patients ([CP, 2/17] versus [VP, 14/19], *p* = 0.0002, Mann-Whitney test) (**Fig. 1A**). Finally, fewer VP recipients had or were receiving an anti-CD20 antibody treatment (6/19; 32%) and were treated with corticosteroids, remdesivir and/or tocilizumab (47% versus 76%) (**Supp. Table 1**).

**Figure 1.**
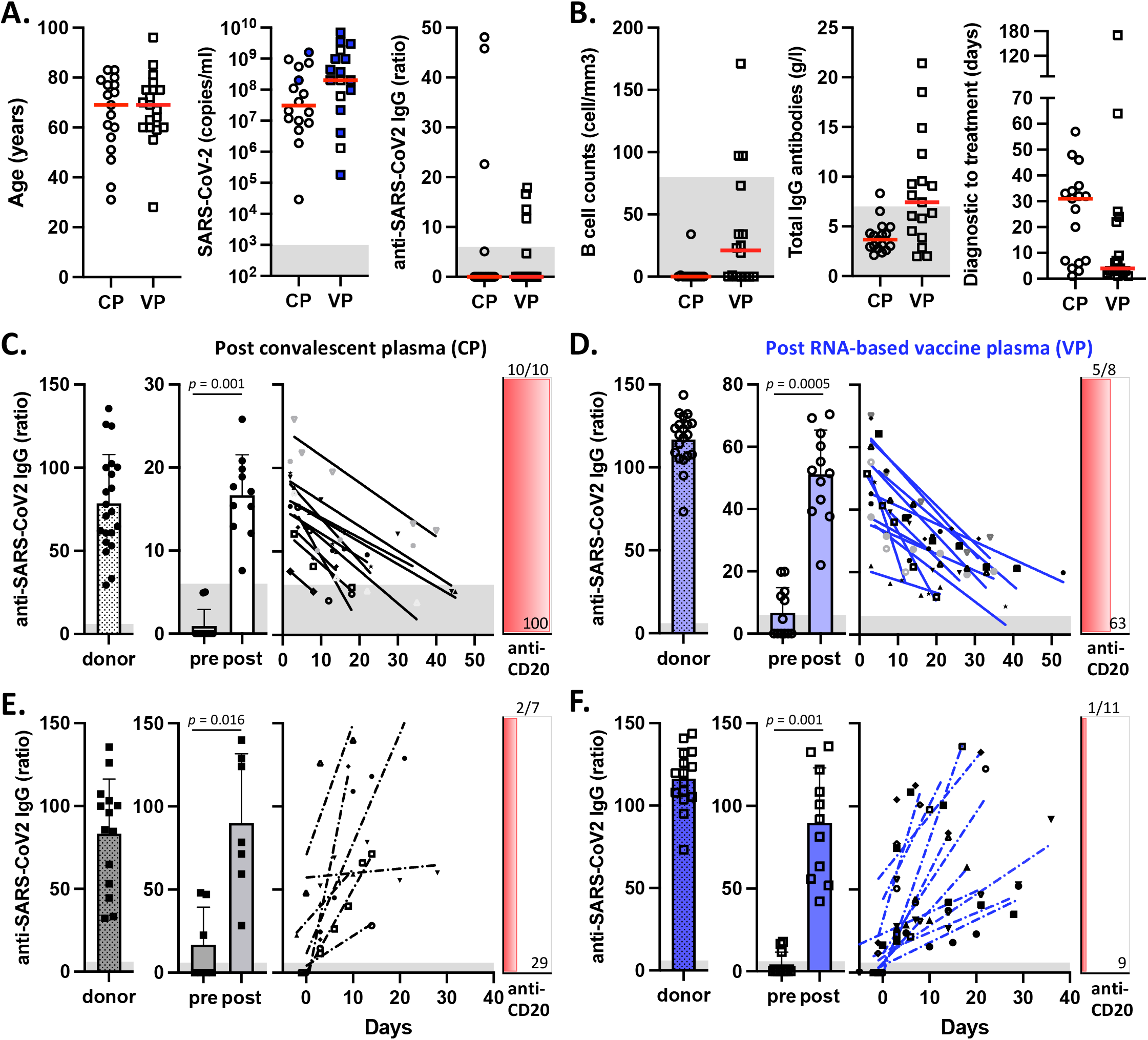
Immune status-related information and serological response follow-up. (**A**) Patient characteristics (age), SARS-CoV-2 viral loads (copies/ml), anti-SARS-CoV-2 S protein IgG antibody levels (ratio) before treatment with convalescent plasma (CP) or vaccinated plasma (VP). Blue symbols represent patients infected by the alpha variant B.1.1.7. (**B**) Absolute B cell counts (cell/mm^3^), total IgG antibody levels (g/l) and time-lapse from diagnostic to treatment (days) before treatment with CP or VP. (**C to F**, left panels) Anti-SARS-CoV-2 S protein IgG titers for each plasma donor post COVID-19 recovery (**C, E**) or after the 2^nd^ injection of an mRNA-based vaccine (**D, F**). (**C to F**, middle panels) Comparison of anti-S IgG antibody titers before and after plasma treatment. The maximum reached value for each patient is depicted. The *p* values are by Wilcoxon matched-pairs signed ranked test. (**C, D**, right panels) Patients presenting anti-S IgG antibody declines following plasma treatment with CP (**C**) or VP (**D**). Of note, anti-S IgG decline kinetic is depicted after each treatment, including those patients who received serial plasma transfusion. (**E, F**, right panels) Patients exhibiting anti-S IgG antibody increases following plasma treatment with CP (**E**) or VP (**F**). (**C to F**, red panels). Proportion of patients (in percentage) pre-exposed to an anti-CD20 antibody targeted treatment.

The majority of patients included in this study had a negative SARS-CoV-2 anti-IgG serology (29/36; 81%) and were not vaccinated (32/36; 89%) (**Fig. 1A**). Most patients had B cell lymphopenia and hypogammaglobulinemia (total IgG) at presentation, both of which were more profound in CP than in VP patients (**Fig. 1B**). At the time of treatment, 11/17 (65%) CP and 8/19 (42%) VP patients needed oxygen supplementation. Among them, 18 patients required high-flow nasal cannula or non-invasive positive pressure ventilation and one patient was intubated on mechanical ventilation support. The median time from diagnosis to plasma treatment was 31 days [1-57 days range] for patients receiving CP treatment. This was reduced to 4 days [1-170 days range] in the cohort of VP, thanks to the systematic diagnostic screening established for every newly-hospitalized patient during the 3^rd^ COVID-19 wave (**Fig. 1B**). One patient had a transient increase in oxygen requirement of unlikely imputability to plasma transfusion, otherwise no transfusion-related adverse events were documented (data not shown). Since we investigated the durable effect of plasma transfusion on the serological and viral evolution of each treated patient, only those patients alive at day 7 after plasma transfusion were included in this study (i.e. 3 patients were censored).

### Anti-CD20 pre-exposition is associated with anti-S IgG titer decay following plasma therapy

B-cell depleting treatment such as anti-CD20 targeted therapies represents an important risk factor for severe forms of COVID-19 (8), which may be related to the failure to mount an efficient endogenous anti-SARS-CoV-2 antibody response. To address this hypothesis, we assessed the anti-S IgG titers of each patient treated either with CP or VP at serial time-points following plasma transfusion. While the antibody levels varied greatly between donors of convalescent plasma, these titers were more homogeneous and in general higher in individuals donating plasma after the 2^nd^ mRNA-based vaccine injection than in unvaccinated convalescent subjects (**Fig. 1C-F**, left panels; donor). This translated into higher levels of anti-S IgG antibodies in patients receiving VP (**Fig. 1C**, middle panel), as compared to those transfused with CP (**Fig. 1D**, middle panel) ([CP, median, ratio of 17] versus [VP, median, ratio of 52], *p* <0.0001, Mann-Whitney test).

Convalescent and vaccinated plasma recipients were further classified according to their antibody kinetic pattern following transfusion. One-half (18/36) of the patients presented a progressive decline in anti-S IgG levels, with a longer time-to-reach negative titers for VP (median, 42 days [15-72]) than for CP (median, 25 days [6-58]; *p* = 0.046, Mann-Whitney test) (**Fig. 1C and D**, right panels). Six of these patients (3 CP and 3 VP), who required additional plasma transfusions due to insufficient clinical and microbiological responses, exhibited an anti-S IgG antibody decline after each treatment. Interestingly, 15 of the 18 patients showing a decline in anti-S IgG levels, had been exposed to an anti-CD20 therapy (10/10 CP patients and 5/8 VP patients). In contrast, the other patients (18/36) showed a progressive increase in anti-S IgG titers following plasma transfusion (**Fig. 1E and F**), and among whom only 3 had received an anti-CD20 treatment (CD20 pre-exposure; [Ab decline, 15/18] versus [Ab increase, 3/18], *p* = 0.0002, Mann-Whitney test). Together, our data indicate that anti-CD20 pre-exposition is associated to a progressive decay in anti-S IgG titers following plasma therapy (CP or VP).

### Patients with progressive decline in anti-S IgG titers following plasma therapy had prolonged SARS-CoV-2 infection before complete virus clearance

At the time of plasma transfusion, patients presented a range from mild to severe COVID-19, according to the WHO classification (CP, mean score at 5, [2 to 9], with 1 mechanically ventilated patient; VP, mean score at 5, [2 to 6]). Three additional patients required mechanical ventilation support upon plasma treatment (2 CP; 1 VP). Clinical improvement in COVID-19 symptoms within a follow-up period of 30 days [13-30 days] after plasma transfusion was reported for 34 of the 36 patients (**Fig. 2A**). Specifically, low WHO scores, between 0 to 1, were attributed for 12/17 (71%) CP patients and 15/19 (79%) VP patients. Moreover, a favorable trend was generally observed for patients who presented an endogenous serological response, compared to those with anti-S IgG antibody declines (**Fig. 2A**). Two patients died from SARS-CoV-2-related complications (1 CP; 1 VP) and 5 (2 CP; 3 VP) from their primary-evolutive malignancy.

**Figure 2.**
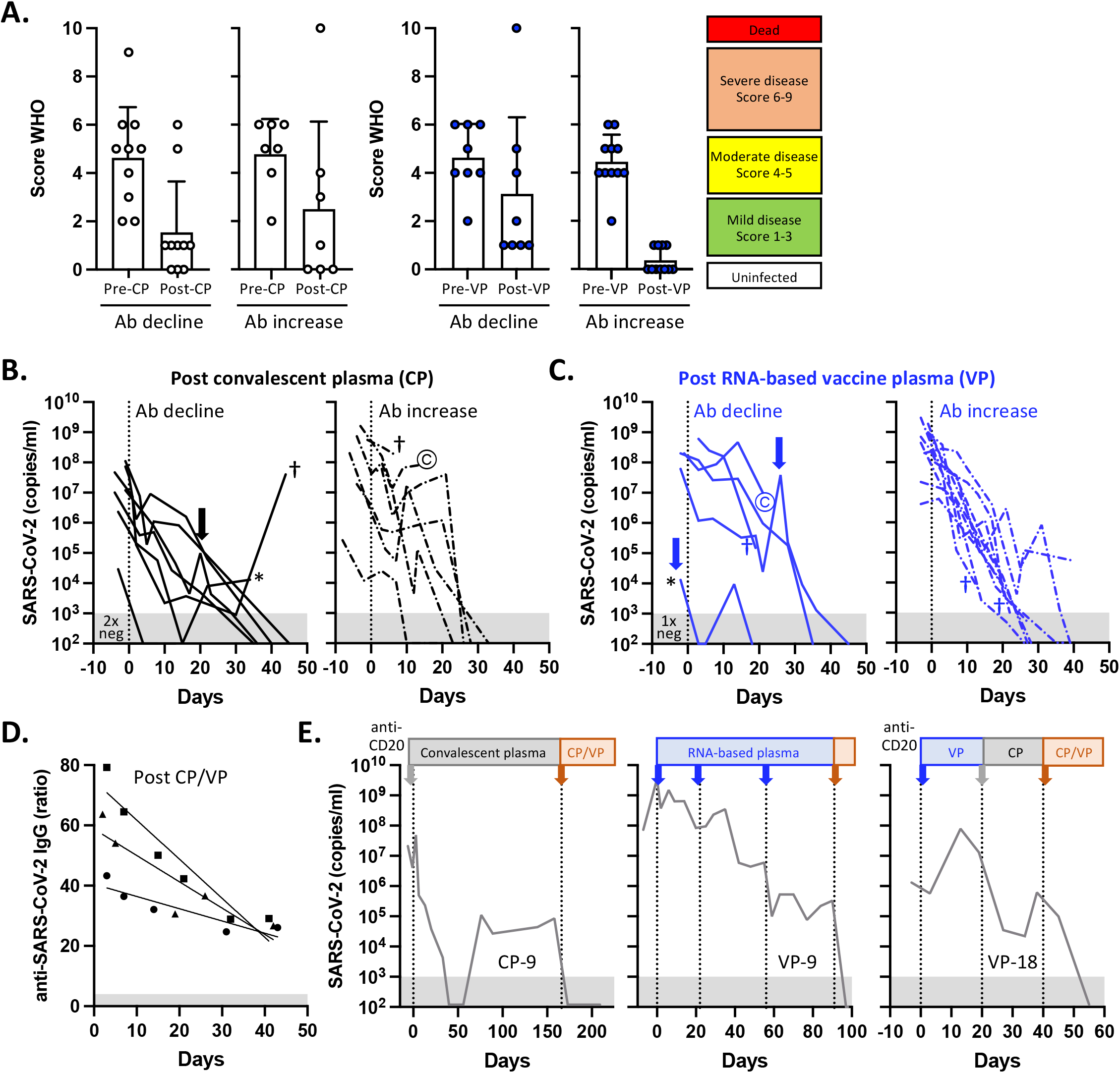
Clinical and viral load recovery in immunocompromised individuals after plasma therapy. (**A**) Clinical status according to the WHO classification before and following CP or VP treatment. Patients were further classified according to their anti-S IgG antibody kinetics (decline versus increase). (**B, C**) Over-time follow-up of SARS-CoV-2 RNA detection in nasopharyngeal swabs (copies/ml) after treatment with CP (**B**) or VP (**C**). Patients were classified according to their anti-S IgG antibody kinetics (decline versus increase). Three patients had undetectable viral loads at D0 of plasma transfusion (2xneg CP, 1xneg VP). Arrows represent patients who received a second plasma treatment. One patient (*) was sequentially treated with CP and VP. Patients who died from SARS-CoV-2 related complications (©) or from their primary-evolutive malignancy (**†**) are depicted. (**D**) Anti-S IgG antibody decline kinetics following convalescent vaccine-boosted plasma treatment (CP/VP, n = 3)). (**E**) Sequential plasma treatments (CP and/or VP and CP/VP) of the 3 patients presenting a prolonged SARS-CoV-2 shedding. (**A-E**) CP, convalescent plasma; VP, vaccinated plasma, CP/VP, convalescent vaccine-boosted plasma.

Alongside, we observed a gradual decline in SARS-CoV-2 RNA levels from nasopharyngeal swabs, with quantitative values ranging below 10E3 copies/ml in 11 (73%) out of 15 CP patients and 13 (72%) of 18 VP patients (**Fig. 2B and C**). Three patients (2 CP, 1 VP) had undetectable viral loads at D0 of plasma transfusion, but still presented clinical and/or radiological signs of active COVID-19. The time-to-negativity was shorter in patients treated with CP or VP, presenting an endogenous anti-SARS-CoV2 response (median, 26 days [13-39]), compared to those with progressive anti-S IgG decline (median 38 days, [4-49], *p* = 0.0032, Mann-Whitney test). Three patients were treated twice, sequentially, either with the same type of plasma or with the other type (CP to VP), allowing for complete viral clearance (**Fig. 2B and C**, see arrows). Finally, three additional patients (CP-9, VP-9, VP-18), who exhibited persistent SARS-CoV-2 shedding, received repeated transfusions (between 2 to 4-times), including plasma from COVID-19 recovered donors boosted by an mRNA vaccine (**Fig. 2D**), leading to the full undetectable SARS-CoV-2 RNA in nasopharyngeal swabs (**Fig. 2E**). Collectively, prolonged SARS-CoV-2 infection was generally observed in the subgroup of patients displaying a progressive decline in anti-S IgG titers following plasma therapy, including the six patients, receiving serial plasma transfusions. With the exception of three patients (VP-7, VP-9 and VP-11), all were pre-treated with anti-CD20.

### Only a minority of patients, unable to mount an endogenous humoral response, presented significant viral evolution following plasma therapy

To investigate whether persistent SARS-CoV-2 infection was associated with intra-host mutation rate following plasma treatment (**Fig. 3**), SARS-CoV-2 genome sequencing was performed on 139 serial respiratory samples from 30 patients, pre- and post-plasma treatment, with a studied interval of up to 182 days (CP, n = 14, [4-182 days]; VP, n = 16, [9-109 days]). Twenty-six out of 30 patients showed one or more intra-host mutations in the viral subpopulations (>10% reads) at any time-point (**Fig. 3A**), some of which reached fixation (>70% reads) over time, supporting the constant within-host virus evolution. Large variations were observed in the number of mutations. Four patients (CP-1, CP-13, VP-3 and VP-4) retained identical viruses over 8 to 22 days, while three others (CP-9, VP-9 and VP-18), who were all unable to mount an endogenous humoral response, presented 26 to 65 mutations, including 20 to 50 that reached fixation for at least one sample time (**Fig. 3A, Supp. Table 2**). Phylogenetic analyses with the most closely-related published genomes from Switzerland supported the monophyletic origin of each strain documented in patients CP-9, VP-9 and VP-18, hence excluding secondary infections with other circulating strains (**Supp. Fig. 1**). Further supporting the persistence of the original virus, CP-9 was infected by B.1.177 until mid-July 2021 and VP-18 by B.1.160 until August 2021, while both variants had disappeared from Switzerland in March 2021, being replaced by B.1.1.7 (alpha variant).

**Figure 3.**
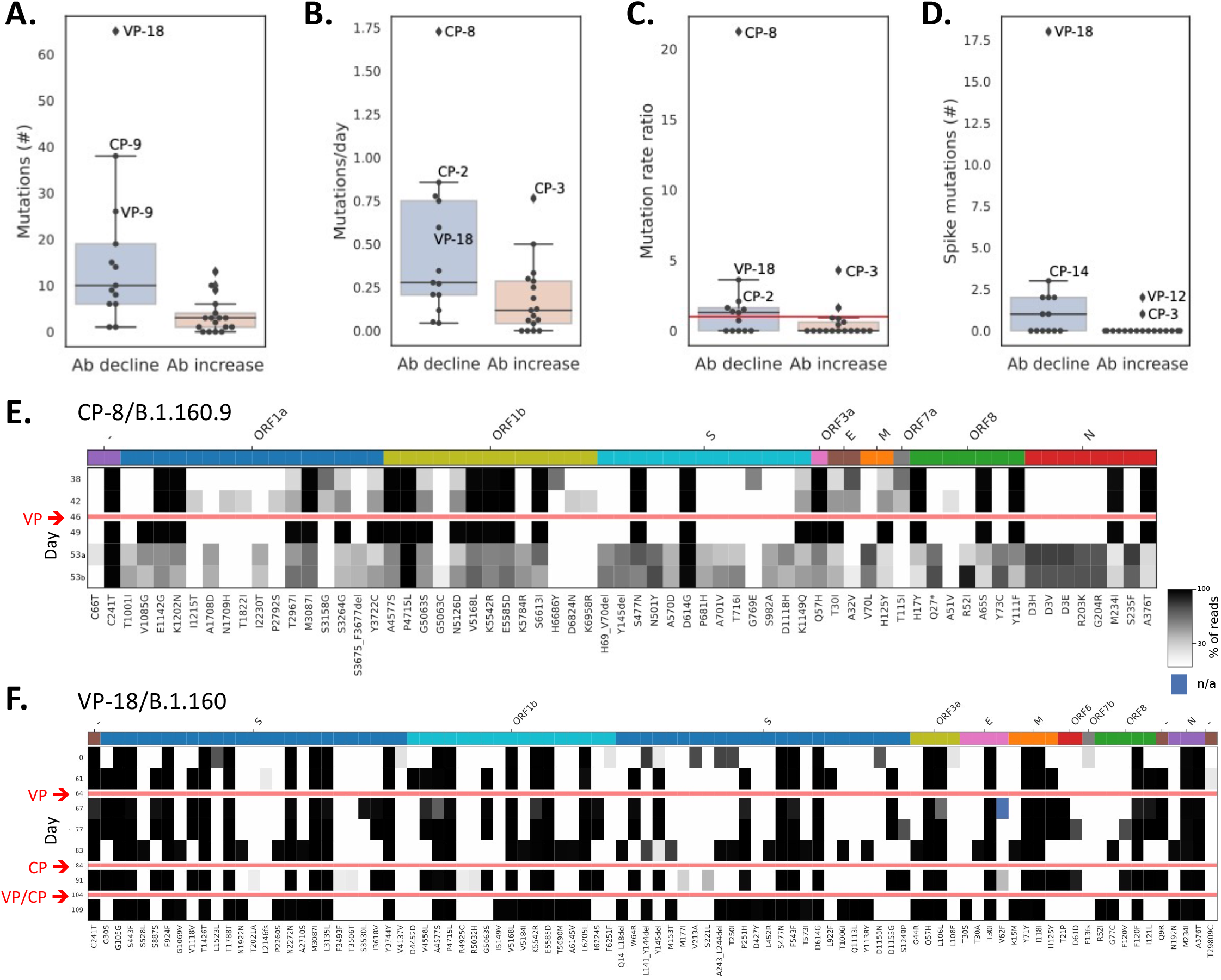
Intra-host viral evolution in immunocompromised patients before and after plasma therapy. **(A)** Number of mutations supported by at least 10% of the reads that differ between sequenced genomes of the same patient. **(B)** Mutation rate calculated as the number of mutations supported by at least 10% of the reads divided by the interval between the first and the last sequenced sample (in days). **(C)** The rate of mutations reaching fixation (>70% of the reads) between the first and last sequenced samples was compared to the theoretical SARS-CoV-2 mutation rate of approximately 25 mutations per year. Patients with a rate ratio larger than one (horizontal red line) present more mutations than expected. **(D)** Number of spike mutations supported by at least 70% of the reads. **(A-D)** Patients were classified according to their anti-S IgG antibody kinetics (decline versus increase). **(E-F)** Overview of identified non-synonymous mutations as compared to the reference Wuhan Hu-1 reference genome, before and/or after plasma therapy for patients CP-8 **(E)** and VP-18 (**F**). Cells indicate the percentage of reads supporting each mutation (rows) in the different samples (columns). Only variants supported by at least 10% of the reads are reported. For CP-8, the sample from day 53 was sequenced twice to rule out a contamination during the sequencing process. CP, convalescent plasma; VP, vaccinated plasma, CP-VP, convalescent vaccine-boosted plasma.

Patients who presented progressive declines in anti-S IgG titers after plasma therapy had significantly higher mutation rates than those showing an endogenous anti-SARS-CoV2 response (**Fig. 3B**, Mann-Whitney U test; *p* = 0.015). Four patients (CP-2, CP-3, CP-8 and VP-18), among which three had declining anti-S IgG titers, presented mutation rates over twice the expected ∼2 mutations per month (**Fig. 3C**) (53). CP-8 presented an exceptionally high number of variants supported by 10-70% of the reads, many of whom reaching fixation in subsequent samples and suggestive of the presence of a heterogeneous viral population (**Fig. 3E**). Viral subpopulations tend to disappear at day 3 following VP treatment, but a very high number of mutations supported by 10-70% of the reads were detected again at day 7. VP-18 also presented an intriguing pattern of mutation acquisition and alternation over time (**Fig. 3F**). In this patient, two distinct mutation profiles were observed alternatively at different time-points (day 61/67/77/91/102 and 83/109), also supported by phylogenetic analyses (**Supp. Fig. 1**). In spite of this viral diversification, consecutive plasma transfusion from different donors, including convalescent vaccine-boosted plasma (CP/V), led to SARS-CoV-2 infection control (**Fig. 2E**).

Only a limited number of spike mutations were globally observed during the follow-up (**Fig. 3D**) and were composed of amino-acid substitutions and several recurrent deletions within the N-terminal domain of the spike protein (e.g. ΔL141-Y144, ΔY145, ΔA243-L244) (**Supp. Table 2**), in line with previous studies (54). Five mutations (S373L, D405N, D427Y, L452R, S494L) were included in the receptor-binding domain (RBD). Patients CP-9 and VP-9 acquired 1 and 2 spike mutations, respectively, at late time-points following plasma treatment (**Supp. Fig. 2 and 3**). Patient VP-18 presented up to 18 spike mutations, among which 10 appeared before plasma treatment (**Supp Table 2, Supp. Fig. 3**). Mutation L452R was acquired on day 83, while ΔL141-Y144, ΔY145 and ΔA243-L244 emerged before plasma treatment (< day 64), yet all four play a potential role as immune escape mutations (54, 55). Patient CP-5 developed 2 novel mutations, among which ΔY145 likely contributes to immune evasion (**Supp. Fig. 2 and 3**) (56). At that time, no second plasma treatment was given, due to the transition to palliative care for her primary evolutive hematological malignancy. Similarly, ΔL141-Y144 deletion, associated to immune resistance after convalescent plasma (54), appeared in another treated patient (CP-14) (**Supp. Fig. 2 and 3**). Overall, our observations indicate that only few immunosuppressed patients presented an accumulation of many mutations over the course of the infection, some of which reached fixation. Patients with progressive anti-S IgG antibody declines following plasma therapy were at highest risk for enhanced viral evolution.

## DISCUSSION

B-cell depleting strategies are broadly used for the treatment of B-cell lymphoid malignancies or inflammatory auto-immune diseases. This study aimed at determining the impact of anti-CD20 monoclonal antibodies (e.g. rituximab, obinutuzumab) on the antibody kinetics and viral evolution upon plasma therapy in hospitalized COVID-19 patients (n = 18/36), in comparison to patients without B-cell depleting treatment (n = 18/36). Patients were transfused with therapeutic plasma from convalescent (CP) or RNA vaccinated (VP) donors. For each patient, a comprehensive longitudinal follow-up (ranging from day 13 up to day 209) combining anti-S protein IgG titer measurements and whole genome sequencing was performed. Our study revealed that the majority (15/18) of patients pre-exposed to anti-CD20 therapies were unable to mount an effective intrinsic humoral response. Nevertheless, 86% of these patients showed viral titer reduction presumably due to the passively transferred neutralizing antibodies. Owing to an insufficient clinical and microbiological response, six of them received additional plasma transfusions but still presented after each treatment, progressive declines in their anti-S IgG titer response. Conversely, a long-term rise in anti-S IgG antibody titers was more frequently observed in non CD20-exposed patients (15/18), and was indicative of an ongoing endogenous response (**Fig. 1**). Overall, 34 of 36 (94%) patients had an improved WHO clinical score within 30 days after plasma therapy and among them, 87% (27/31, excluding 3 patients with undetectable viral loads before plasma transfusion) presented no SARS-CoV-2 RNA detection by PCR (**Fig. 2**). Our results confirm the efficacy of plasma therapy in this setting and may further support the key role of donor plasma selected for enhanced anti-SARS-CoV-2 antibody titers in correcting humoral deficiency and improving clinical outcomes for patients with B-cell depleting immunotherapy, in line with previous reports, (3, 57).

The effectiveness of CP is likely influenced by the quantity of neutralizing antibodies (correlating to the titers of anti-S IgG antibodies) present at the time of donation (13, 58). Initially, Libster and coworkers (12) reported a dose-dependent effect relative to the antibody titers after transfusion, with reduced COVID-19 progression. In early 2021, initiation of the Swiss vaccine campaign further enabled collecting non-COVID-19 donor plasma enriched with high and homogenous anti-S protein IgG titers post-second mRNA vaccination (**Fig. 1**) (i.e. with high neutralizing titers (40, 41)). In the meantime, two doses of mRNA vaccines were shown to remain highly effective against symptomatic SARS-CoV-2 infection and severe outcomes with different variants of concern (59, 60). Moreover, a single immunization can boost the neutralizing titers up to 1000-fold in COVID-19-recovered donors (61). When administrated to immunosuppressed patients, comprising those who received an anti-CD20 pre-treatment, vaccinated plasma allowed the efficient transfer of anti-S IgG antibodies and led to clinical and viral load recovery comparable to CP therapy (**Fig. 1 and 2**). In addition, three cases presenting persistent SARS-CoV-2 infection were efficiently treated with convalescent vaccine-boosted plasma (**Fig. 2**). Collectively, our data show that vaccine-based plasma may represent an alternative treatment alongside to convalescent plasma, in the management of COVID-19 patients with B-cell lymphopenia. Besides, the use of plasma from vaccine-boosted convalescent individuals or from vaccinated ones boosted by a breakthrough infection likely broadens the spectrum of anti-SARS-CoV-2 humoral response, especially against variants such as Omicron to which convalescent-or vaccinated-only donors have not been exposed (61-63) and is currently the only source of plasma.

Only a minority of the 30 genotyped patients displayed an increased viral mutation rate (**Fig. 3**), most of whom were unable to mount an intrinsic antibody response to SARS-CoV-2. Likewise, whole-genome sequencing showed the emergence of a limited number of spike mutations (e.g. ΔL141-Y144, ΔY145 and L452R) potentially associated to immune escape, in different patients (CP-14, CP-5 and VP-18, respectively) following plasma therapy (**Fig. 3**). Patient VP-18 was exceptional as he presented many fixed mutations (20/52) after the initial two months of infection, before plasma therapy. Moreover, the alternation of two mutation patterns suggested the selection of undetected virus subpopulations at different time-points possibly related to the serial plasma therapies administrated to this patient.

Thorough follow-up allowed identifying a few cases with prolonged viral shedding who needed serial transfusion for a complete recovery. Our data is in line with a systematic review by Focosi *et al*. (54), reporting that convalescent plasma may be associated to a lower risk of emergence of resistant variants, contrasting with the documented immune escape after treatment with monoclonal Abs. This may in part be explained by the polyclonal nature of the transfused anti-SARS-CoV-2 antibodies. Moreover, escape variants associated to plasma therapy exhibited recurrent amino acid deletions in the NTD region as well as single amino acid changes throughout the spike protein (**Supp. Table 2**). Consistently, only 5 of the 31 identified spike mutations affected the RBD region. In line with these observations, escape from polyclonal plasmas likely involves larger antigenic structural changes than escape from monoclonal Abs, targeting single epitopes (54). As comparable viral evolution patterns were found following transfusion with vaccine-based plasma, this also suggests that convalescent and vaccinated plasma may share common mechanisms in antibody-mediated protection.

In summary, our case series extends on previous findings (3, 57), validating the concept that immunosuppressed patients, particularly those who are pre-exposed to an anti-CD20 monoclonal antibody treatment, are unable to produce a potent anti-SARS-CoV-2 humoral response and rely on the passive transfer of neutralizing antibodies. Such immunosuppressed patients with a *de novo* SARS-CoV-2 infection should quickly be identified at the daily clinical practice, since most of them require individualized clinical care and follow-up. Moreover, B cell-depleted COVID-19 patients are at increased risk for long-term viral replication, as compared to other vulnerable individuals who are still able to develop their own endogenous antibody response. High mutation rate was only observed in few patients with prolonged virus shedding. Yet, this observation emphasizes the need for long-term surveillance for the emergence of new variants carrying mutations favoring escape to current population immunity by regular SARS-CoV-2 viral load and genomic monitoring. Finally, given the importance of the humoral immune response for clinical recovery (11), plasma therapy from convalescent vaccine-boosted donors remains a rational option, since it is unexpensive and logistically easy to organize, and contains high titers of neutralizing antibodies against SARS-CoV-2 associated to a broad antigenic spectrum (61-63).

## Supporting information

Supplemental Informations

## Data Availability

All data produced in the present study are available upon reasonable request to the authors

## AUTHOR CONTRIBUTIONS

Study design and writing of the manuscript: DG, TP, NR. Patient care and acquisition of data: DG, TG, DB, DD, PV, OM, NR. Analysis and interpretation of data: DG, TP, TG, GG, CB, NR. Revision and approval of the manuscript: DG, TP, TG, OO, SF, PV, OM, GG, CB, NR.

## ACKNOWLEDGMENTS

We are grateful to the patients who consented to the anonymous use of their clinical data for this study and the blood donors for their dedicated collaboration in this study. We would like to acknowledge all the clinicians for their participation in data collection: S. Frascarolo, M. Monti, W. Bosshard, B. Guery, R. Stadelmann, and the participating medical and laboratory teams of the Lausanne University Hospital (Lausanne, Switzerland). We thank H. Andreu-Ullrich, B. Banwarth, S. Berger, M. Burgener, G. Canellini, J. Conne, C. Corisello, D. Crettaz, N. Doegnitz, K. Eichler, M. Finger, E. Gagliarducci, M. Gavillet, M. Graziani, D. Huber Marcantonio, M-L. Métrailler, V. Morin, C. Niederhauser, M. Prudent, C. Tinguely, and I. Weigand, as well as the blood center teams of the Interregional Blood Transfusion SRC (Bern-Vaud-Valais, Switzerland). We are thankful to Prof. Daniel E. Speiser for comments and critical reading of the manuscript.

## SUPPLEMENTARY INFORMATION

Complementary clinical and methodological information for this case report can be found in the **Supplementary Information**.

## REFERENCES

1. Williamson EJ, Walker AJ, Bhaskaran K, Bacon S, Bates C, Morton CE, et al. Factors associated with COVID-19-related death using OpenSAFELY. Nature. 2020;584(7821):430–6.

2. Vijenthira A, Gong IY, Fox TA, Booth S, Cook G, Fattizzo B, et al. Outcomes of patients with hematologic malignancies and COVID-19: a systematic review and meta-analysis of 3377 patients. Blood. 2020;136(25):2881–92.

3. Thompson MA, Henderson JP, Shah PK, Rubinstein SM, Joyner MJ, Choueiri TK, et al. Association of Convalescent Plasma Therapy With Survival in Patients With Hematologic Cancers and COVID-19. JAMA Oncol. 2021.

4. Pagano L, Salmanton-Garcia J, Marchesi F, Busca A, Corradini P, Hoenigl M, et al. COVID-19 infection in adult patients with hematological malignancies: a European Hematology Association Survey (EPICOVIDEHA). J Hematol Oncol. 2021;14(1):168.

5. Russell B, Moss CL, Shah V, Ko TK, Palmer K, Sylva R, et al. Risk of COVID-19 death in cancer patients: an analysis from Guy’s Cancer Centre and King’s College Hospital in London. Br J Cancer. 2021;125(7):939–47.

6. Avouac J, Drumez E, Hachulla E, Seror R, Georgin-Lavialle S, El Mahou S, et al. COVID-19 outcomes in patients with inflammatory rheumatic and musculoskeletal diseases treated with rituximab: a cohort study. Lancet Rheumatol. 2021;3(6):e419–e26.

7. Tepasse PR, Hafezi W, Lutz M, Kuhn J, Wilms C, Wiewrodt R, et al. Persisting SARS-CoV-2 viraemia after rituximab therapy: two cases with fatal outcome and a review of the literature. Br J Haematol. 2020;190(2):185–8.

8. Dulery R, Lamure S, Delord M, Di Blasi R, Chauchet A, Hueso T, et al. Prolonged in-hospital stay and higher mortality after Covid-19 among patients with non-Hodgkin lymphoma treated with B-cell depleting immunotherapy. Am J Hematol. 2021;96(8):934–44.

9. Thakkar A, Gonzalez-Lugo JD, Goradia N, Gali R, Shapiro LC, Pradhan K, et al. Seroconversion rates following COVID-19 vaccination among patients with cancer. Cancer Cell. 2021;39(8):1081–90 e2.

10. Cattaneo C, Cancelli V, Imberti L, Dobbs K, Sottini A, Pagani C, et al. Production and persistence of specific antibodies in COVID-19 patients with hematologic malignancies: role of rituximab. Blood Cancer J. 2021;11(9):151.

11. Seebacher NA. The antibody response of haematological malignancies to COVID-19 infection and vaccination. Br J Cancer. 2022.

12. Libster R, Perez Marc G, Wappner D, Coviello S, Bianchi A, Braem V, et al. Early High-Titer Plasma Therapy to Prevent Severe Covid-19 in Older Adults. N Engl J Med. 2021;384(7):610–8.

13. Joyner MJ, Carter RE, Senefeld JW, Klassen SA, Mills JR, Johnson PW, et al. Convalescent Plasma Antibody Levels and the Risk of Death from Covid-19. N Engl J Med. 2021;384(11):1015–27.

14. Simonovich VA, Burgos Pratx LD, Scibona P, Beruto MV, Vallone MG, Vazquez C, et al. A Randomized Trial of Convalescent Plasma in Covid-19 Severe Pneumonia. N Engl J Med. 2021;384(7):619–29.

15. Menichetti F, Popoli P, Puopolo M, Spila Alegiani S, Tiseo G, Bartoloni A, et al. Effect of High-Titer Convalescent Plasma on Progression to Severe Respiratory Failure or Death in Hospitalized Patients With COVID-19 Pneumonia: A Randomized Clinical Trial. JAMA Netw Open. 2021;4(11):e2136246.

16. Group RC. Convalescent plasma in patients admitted to hospital with COVID-19 (RECOVERY): a randomised controlled, open-label, platform trial. Lancet. 2021;397(10289):2049–59.

17. Begin P, Callum J, Jamula E, Cook R, Heddle NM, Tinmouth A, et al. Convalescent plasma for hospitalized patients with COVID-19: an open-label, randomized controlled trial. Nat Med. 2021;27(11):2012–24.

18. Writing Committee for the R-CAPI, Estcourt LJ, Turgeon AF, McQuilten ZK, McVerry BJ, Al-Beidh F, et al. Effect of Convalescent Plasma on Organ Support-Free Days in Critically Ill Patients With COVID-19: A Randomized Clinical Trial. JAMA. 2021;326(17):1690–702.

19. Weinreich DM, Sivapalasingam S, Norton T, Ali S, Gao H, Bhore R, et al. REGN-COV2, a Neutralizing Antibody Cocktail, in Outpatients with Covid-19. N Engl J Med. 2021;384(3):238–51.

20. Hueso T, Pouderoux C, Pere H, Beaumont AL, Raillon LA, Ader F, et al. Convalescent plasma therapy for B-cell-depleted patients with protracted COVID-19. Blood. 2020;136(20):2290–5.

21. Tremblay D, Seah C, Schneider T, Bhalla S, Feld J, Naymagon L, et al. Convalescent Plasma for the Treatment of Severe COVID-19 Infection in Cancer Patients. Cancer Med. 2020;9(22):8571–8.

22. Ljungquist O, Lundgren M, Iliachenko E, Mansson F, Bottiger B, Landin-Olsson M, et al. Convalescent plasma treatment in severely immunosuppressed patients hospitalized with COVID-19: an observational study of 28 cases. Infect Dis (Lond). 2021:1–9.

23. Lang-Meli J, Fuchs J, Mathe P, Ho HE, Kern L, Jaki L, et al. Case Series: Convalescent Plasma Therapy for Patients with COVID-19 and Primary Antibody Deficiency. J Clin Immunol. 2021.

24. Rodionov RN, Biener A, Spieth P, Achleitner M, Holig K, Aringer M, et al. Potential benefit of convalescent plasma transfusions in immunocompromised patients with COVID-19. Lancet Microbe. 2021;2(4):e138.

25. Kenig A, Ishay Y, Kharouf F, and Rubin L. Treatment of B-cell depleted COVID-19 patients with convalescent plasma and plasma-based products. Clin Immunol. 2021;227:108723.

26. Focosi D, and Franchini M. Potential use of convalescent plasma for SARS-CoV-2 prophylaxis and treatment in immunocompromised and vulnerable populations. Expert Rev Vaccines. 2021:1–8.

27. Senefeld JW, Klassen SA, Ford SK, Senese KA, Wiggins CC, Bostrom BC, et al. Use of convalescent plasma in COVID-19 patients with immunosuppression. Transfusion. 2021;61(8):2503–11.

28. Corey L, Beyrer C, Cohen MS, Michael NL, Bedford T, and Rolland M. SARS-CoV-2 Variants in Patients with Immunosuppression. N Engl J Med. 2021;385(6):562–6.

29. Aydillo T, Gonzalez-Reiche AS, Aslam S, van de Guchte A, Khan Z, Obla A, et al. Shedding of Viable SARS-CoV-2 after Immunosuppressive Therapy for Cancer. N Engl J Med. 2020;383(26):2586–8.

30. Choi B, Choudhary MC, Regan J, Sparks JA, Padera RF, Qiu X, et al. Persistence and Evolution of SARS-CoV-2 in an Immunocompromised Host. N Engl J Med. 2020;383(23):2291–3.

31. Avanzato VA, Matson MJ, Seifert SN, Pryce R, Williamson BN, Anzick SL, et al. Case Study: Prolonged Infectious SARS-CoV-2 Shedding from an Asymptomatic Immunocompromised Individual with Cancer. Cell. 2020;183(7):1901–12 e9.

32. Kemp SA, Collier DA, Datir RP, Ferreira I, Gayed S, Jahun A, et al. SARS-CoV-2 evolution during treatment of chronic infection. Nature. 2021;592(7853):277–82.

33. Truong TT, Ryutov A, Pandey U, Yee R, Goldberg L, Bhojwani D, et al. Increased viral variants in children and young adults with impaired humoral immunity and persistent SARS-CoV-2 infection: A consecutive case series. EBioMedicine. 2021;67:103355.

34. Borges V, Isidro J, Cunha M, Cochicho D, Martins L, Banha L, et al. Long-Term Evolution of SARS-CoV-2 in an Immunocompromised Patient with Non-Hodgkin Lymphoma. mSphere. 2021;6(4):e0024421.

35. Andreano E, Piccini G, Licastro D, Casalino L, Johnson NV, Paciello I, et al. SARS-CoV-2 escape from a highly neutralizing COVID-19 convalescent plasma. Proc Natl Acad Sci U S A. 2021;118(36).

36. Honjo K, Russell RM, Li R, Liu W, Stoltz R, Tabengwa EM, et al. Convalescent plasma-mediated resolution of COVID-19 in a patient with humoral immunodeficiency. Cell Rep Med. 2021;2(1):100164.

37. Long QX, Liu BZ, Deng HJ, Wu GC, Deng K, Chen YK, et al. Antibody responses to SARS-CoV-2 in patients with COVID-19. Nat Med. 2020;26(6):845–8.

38. Jabal KA, Wiegler KB, and Edelstein M. Convalescent plasma from people vaccinated after COVID-19 infection. Lancet Microbe. 2021;2(5):e171–e2.

39. Katz LM. (A Little) Clarity on Convalescent Plasma for Covid-19. N Engl J Med. 2021;384(7):666–8.

40. Walsh EE, Frenck RW, Jr., Falsey AR, Kitchin N, Absalon J, Gurtman A, et al. Safety and Immunogenicity of Two RNA-Based Covid-19 Vaccine Candidates. N Engl J Med. 2020;383(25):2439–50.

41. Jackson LA, Anderson EJ, Rouphael NG, Roberts PC, Makhene M, Coler RN, et al. An mRNA Vaccine against SARS-CoV-2 - Preliminary Report. N Engl J Med. 2020;383(20):1920–31.

42. Fenwick C, Croxatto A, Coste AT, Pojer F, Andre C, Pellaton C, et al. Changes in SARS-CoV-2 Spike versus Nucleoprotein Antibody Responses Impact the Estimates of Infections in Population-Based Seroprevalence Studies. J Virol. 2021;95(3).

43. Fenwick C, Turelli P, Pellaton C, Farina A, Campos J, Raclot C, et al. A high-throughput cell-and virus-free assay shows reduced neutralization of SARS-CoV-2 variants by COVID-19 convalescent plasma. Sci Transl Med. 2021;13(605).

44. Zimmerli A, Monti M, Fenwick C, Eckerle I, Beigelman-Aubry C, Pellaton C, et al. Case Report: Stepwise Anti-Inflammatory and Anti-SARS-CoV-2 Effects Following Convalescent Plasma Therapy With Full Clinical Recovery. Front Immunol. 2021;12:613502.

45. Opota O, Brouillet R, Greub G, and Jaton K. Comparison of SARS-CoV-2 RT-PCR on a high-throughput molecular diagnostic platform and the cobas SARS-CoV-2 test for the diagnostic of COVID-19 on various clinical samples. Pathog Dis. 2020;78(8).

46. Jacot D, Greub G, Jaton K, and Opota O. Viral load of SARS-CoV-2 across patients and compared to other respiratory viruses. Microbes Infect. 2020;22(10):617–21.

47. Kubik S, Marques AC, Xing X, Silvery J, Bertelli C, De Maio F, et al. Recommendations for accurate genotyping of SARS-CoV-2 using amplicon-based sequencing of clinical samples. Clin Microbiol Infect. 2021;27(7):1036 e1–e8.

48. Garrison E, Marth G. Haplotype-based variant detection from short-read sequencing. arXiv preprint 1207.3907 [q-bio.GN] 2012.

49. Danecek P, Bonfield JK, Liddle J, Marshall J, Ohan V, Pollard MO, et al. Twelve years of SAMtools and BCFtools. Gigascience. 2021;10(2).

50. Jacot D, Pillonel T, Greub G, and Bertelli C. Assessment of SARS-CoV-2 Genome Sequencing: Quality Criteria and Low-Frequency Variants. J Clin Microbiol. 2021;59(10):e0094421.

51. O’Toole A, Scher E, Underwood A, Jackson B, Hill V, McCrone JT, et al. Assignment of epidemiological lineages in an emerging pandemic using the pangolin tool. Virus Evol. 2021;7(2):veab064.

52. Hadfield J, Megill C, Bell SM, Huddleston J, Potter B, Callender C, et al. Nextstrain: real-time tracking of pathogen evolution. Bioinformatics. 2018;34(23):4121–3.

53. Duchene S, Featherstone L, Haritopoulou-Sinanidou M, Rambaut A, Lemey P, and Baele G. Temporal signal and the phylodynamic threshold of SARS-CoV-2. Virus Evol. 2020;6(2):veaa061.

54. Focosi D, Maggi F, Franchini M, McConnell S, and Casadevall A. Analysis of Immune Escape Variants from Antibody-Based Therapeutics against COVID-19: A Systematic Review. Int J Mol Sci. 2021;23(1).

55. Alkhatib M, Svicher V, Salpini R, Ambrosio FA, Bellocchi MC, Carioti L, et al. SARS-CoV-2 Variants and Their Relevant Mutational Profiles: Update Summer 2021. Microbiol Spectr. 2021;9(3):e0109621.

56. McCallum M, De Marco A, Lempp FA, Tortorici MA, Pinto D, Walls AC, et al. N-terminal domain antigenic mapping reveals a site of vulnerability for SARS-CoV-2. Cell. 2021;184(9):2332–47 e16.

57. Hueso T, Godron AS, Lanoy E, Pacanowski J, Levi LI, Gras E, et al. Convalescent plasma improves overall survival in patients with B-cell lymphoid malignancy and COVID-19: a longitudinal cohort and propensity score analysis. Leukemia. 2022.

58. Korper S, Jahrsdorfer B, Corman VM, Pilch J, Wuchter P, Blasczyk R, et al. Donors for SARS-CoV-2 Convalescent Plasma for a Controlled Clinical Trial: Donor Characteristics, Content and Time Course of SARS-CoV-2 Neutralizing Antibodies. Transfus Med Hemother. 2021;48(3):137–47.

59. Lopez Bernal J, Andrews N, Gower C, Gallagher E, Simmons R, Thelwall S, et al. Effectiveness of Covid-19 Vaccines against the B.1.617.2 (Delta) Variant. N Engl J Med. 2021;385(7):585–94.

60. Nasreen S, Chung H, He S, Brown KA, Gubbay JB, Buchan SA, et al. Effectiveness of COVID-19 vaccines against symptomatic SARS-CoV-2 infection and severe outcomes with variants of concern in Ontario. Nat Microbiol. 2022.

61. Stamatatos L, Czartoski J, Wan YH, Homad LJ, Rubin V, Glantz H, et al. mRNA vaccination boosts cross-variant neutralizing antibodies elicited by SARS-CoV-2 infection. Science. 2021.

62. Evans JP, Zeng C, Carlin C, Lozanski G, Saif LJ, Oltz EM, et al. Neutralizing antibody responses elicited by SARS-CoV-2 mRNA vaccination wane over time and are boosted by breakthrough infection. Sci Transl Med. 2022:eabn8057.

63. Jahrsdorfer B, Fabricius D, Scholz J, Ludwig C, Grempels A, Lotfi R, et al. BNT162b2 Vaccination Elicits Strong Serological Immune Responses Against SARS-CoV-2 Including Variants of Concern in Elderly Convalescents. Front Immunol. 2021;12:743422.

