## Supplemental Informations for "Antibody response and intra-host viral evolution after plasma therapy in COVID-19 patients pre-exposed or not to B-cell depleting agents"

### SUPPLEMENTAL FIGURE LEGENDS

**Supp. Fig. 1. Phylogenetic analysis of three immunocompromised patients with prolonged SARS-CoV-2 infection.** Maximum likelihood phylogenetic tree obtained from patient sequences along with a set of representative SARS-CoV-2 genome sequences from Switzerland between September 2020 and February 2021. (**A**) Patient VP-18 was infected with a virus from lineage B.1.160 that accumulated 65 mutations in 109 days. (**B**) Patient VP-9 was infected with a virus from lineage B.1.1.7 that accumulated 26 mutations over 96 days (**C**) Patient CP-9 was infected by a lineage B.1.177 variant that accumulated 38 mutation over 182 days. All three phylogenies support a common origin of each patient sequenced SARS-CoV-2 genomes.

**Supp. Fig. 2. Longitudinal assessment of viral evolution in immunocompromised patients before and after plasma therapy.** SARS-CoV-2 whole-genome viral sequencing was performed on longitudinally collected nasopharyngeal swaps from CP versus VP treated patients. **(A-D)** Overview of identified non-synonymous mutations as compared to the reference Wuhan Hu-1 reference genome, before and/or after plasma therapy for patients CP-9 **(A)**, VP-9 **(B)**, CP-5 **(C)** and CP-14 **(D)**. Cells indicate the percentage of reads supporting each mutation (rows) in the different samples (columns). Only variants supported by at least 10% of the reads are reported. Of note, patient CP-9, VP-9 and CP-14 received serial plasma treatment. CP, convalescent plasma; VP, vaccinated plasma, CP/VP, convalescent vaccine-boosted plasma.

**Supp. Fig. 3. Longitudinal assessment of spike mutations in immunocompromised patients before and after plasma therapy. (A-E)** Overview of identified non-synonymous mutations in the spike protein as compared to the reference Wuhan Hu-1 reference genome, before and/or after plasma therapy for patients CP-9 **(A)**, CP-5 **(B)**, CP-14 **(C),** VP-9 **(D)**, and VP-18 **(E)**. Cells indicate the percentage of reads supporting each mutation (rows) in the different samples (columns). Only variants supported by at least 10% of the reads are reported. Of note, patient CP-9, CP-14, VP-9 and VP-18 received serial plasma treatment. CP, convalescent plasma; VP, vaccinated plasma, CP-VP, convalescent vaccine-boosted plasma.

### Supplemental Table 1. Patient characteristics

| | Convalescent plasma (n = 17) | Vaccine-based plasma (n = 19) |
|---|---|---|
| **Females/males ratio, n** | 6/11 | 9/10 |
| **Nosocomial infection, n (%)** | 2/17 (12) | 12/19 (63) |
| **Hematological malignancies, n (%)** | 15/17 (88) | 7/19 (37) |
| Diffuse large B-cell lymphoma | 4 | 1 |
| Follicular lymphoma | 3 | 2 |
| Mantle cell lymphoma | 1 | 1 |
| Posttransplant lymphoproliferative disorder (PTLD) [1] | 2* | - |
| Multiple myeloma | 2 | - |
| Amyloidosis | - | 1 |
| Waldenström macroglobulinemia | 1 | - |
| Small lymphocytic lymphoma | 1 | - |
| Angioimmunoblastic T-cell lymphoma | - | 1 |
| Acute T-lymphoblastic leukemia | 1 | - |
| Acute myeloid leukemia | - | 1 |
| **Non-hematological diseases, n (%)** | 4/17 (24) | 9/19 (47) |
| Autoimmune disease [2] | 2 | 3 |
| Solid organ transplant [3] | 2* | 3[#] |
| Solid tumor [4] | - | 3[#] |
| **Other high-risk patients, n (%) [5]** | - | 4/19 (21) |
| **Hematological disease status, n** | | |
| At diagnosis | 2 | 2 |
| Complete response | 8 | 4 |
| Partial response | 2 | - |
| Progressive disease | 3 | 1 |
| **Immunosuppressive treatments** | | |
| Rituximab, other anti-CD20 treatments [6] | 12/17 (71) | 6/19 (31.5) |
| - Past | 6 | 1 |
| - Ongoing | 6 | 5 |
| Other immunosuppressive treatments [7] | 6/17 (35) | 7/19 (37) |
| Not attributed | - | 6/19 (31.5) |
| **COVID-19 related treatments, n (%)** | 13/17 (76) | 9/19 (47) |
| Steroids | 12 | 9 |
| Remdesivir | 4 | 2 |
| Tocilizumab | 1 | - |
| **Survival, n (%)** | 14/17 (82) | 15/19 (79) |
| COVID-related death | 1 | 1 |
| Primary disease-related death | 2 | 3 |

[1] PTLD subtypes: Burkitt lymphoma (1), primary effusion lymphoma (1), resulting from solid organ transplant (*); [2] Rheumatoid arthritis (3), multiple sclerosis (1), granulomatosis with polyangiitis (1); [3] Kidney (3), pancreatic (1) and cardiac (1) transplants; [4] Small bowel adenocarcinoma (1) in a kidney transplant patient [#], intraductal papillary and mucinous pancreatic tumor (1), esophageal small-cell neuroendocrine carcinoma (1); [5] High-risk patients with nosocomial infection <72h post symptoms or diagnosis; [6] Alone or in association with Venetoclax (1), methotrexate and procarbazin followed by BCNU-thiotepa intensification and autologous stem cell transplantation (ASCT) (1), CHOP (cyclophosphamide, doxorubicin, vincristine and prednisone) alternating high-dose methotrexate (1), CHOP followed by BEAM (BCNU + Etoposide + Cytarabine + Melphalan) intensification and ASCT (1); CHOP (4), Obinutuzumab-Lenalidomid (1), Bendamustin (4); [7] Induction regimen with FLAG (fludarabine + high-dose cytarabine + G-CSF)-idarubicin-venetoclax (1) or with high-dose cytarabine, daunorubicin and Venetoclax (1), tandem intensification by melphalan and ASCT (1), Venetoclax monotherapy (1), Daratumumab monotherapy (1), plasmapheresis followed by Bendamustin monotherapy (1), Isatuximab, pomalidomide and dexamethasone (1), Ibrutinib monotherapy (1), Methotrexate monotherapy (1), chronic treatment with prednisone and cyclosporine (1), CHOP (1), chronic treatment with mycophenolate mofetil and tacrolimus (2), FOLFIRI (folinic acid, fluorouracil, irinotecan) (1).

### Supplemental Table 2. Intra-host variations before and/or after plasma therapy

| Patient code[1] | Sex | Anti-CD20 | N° of plasma TT | Strain | Interval days | N° of genomes | Variable mutations[2] | Mutation rate ratio[3] | Mutations in spike domain[4] |
|---|---|---|---|---|---|---|---|---|---|
| CP-2 | M | yes | 1x | B.1.160 | 7 | 4 | 6 | 2.09 | - |
| CP-4 | F | yes | 1x | B.1.1.269 | 20 | 5 | 15 | 0 | - |
| CP-5 | F | yes | 1x | B.1.258 | 68 | 4 | 8 | 1.29 | ΔY145, S494L |
| CP-7 | F | yes | 2x | B.1.177 | 18 | 6 | 14 | 0 | - |
| CP-8 | M | yes | 1x | B.1.160.9 | 11 | 3 | 19 | 21.24 | K1149Q |
| CP-9 | F | yes | 2x | B.1.177 | 182 | 7 | 38 | 0.72 | A67V |
| CP-14 | M | yes | 2x | B.1.160 | 29 | 3 | 6 | 1.51 | T19A, ΔL141-Y144, S373L |
| CP-1 | M | no | 1x | B.1.177.44 | 22 | 3 | 0 | 0 | - |
| CP-3 | M | yes | 1x | B.1.177.44 | 17 | 2 | 13 | 4.29 | P9L |
| CP-10 | F | yes | 1x | B.1.416.1 | 24 | 2 | 1 | 0 | - |
| CP-11 | M | no | 1x | B.1.258.17 | 20 | 3 | 10 | 0 | - |
| CP-13 | M | no | 1x | B.1.160 | 8 | 2 | 0 | 0 | - |
| CP-15 | M | no | 1x | B.1.1.7 | 17 | 3 | 2 | 0 | - |
| CP-16 | F | no | 1x | B.1.1.7 | 24 | 4 | 3 | 0.61 | - |
| VP-1 | M | yes | 1x | B.1.1.7 | 23 | 2 | 1 | 0 | - |
| VP-2 | F | yes | 2x | B.1.221 | 36 | 7 | 10 | 1.62 | P25T, A97T |
| VP-7 | M | no | 1x | B.1.1.7 | 20 | 6 | 1 | 0 | - |
| VP-9 | M | no | 4x | B.1.1.7 | 96 | 18 | 26 | 1.37 | S12F, P174S |
| VP-11 | M | no | 1x | B.1.1.7 | 26 | 6 | 9 | 0 | - |
| VP-18 | M | yes | 3x | B.1.160 | 109 | 7 | 65 | 3.62 | ΔQ14-L18, W64R, ΔL141-Y144, ΔY145, M153T, V213A, ΔA243-L244, T250I, P251H, D427Y, L452R, T573I, L922F, T1006I, Q1113L, D1153G, D1153N, S1249P |
| VP-3 | F | no | 1x | B.1.1.7 | 16 | 2 | 0 | 0 | - |
| VP-4 | F | no | 1x | B.1.1.7 | 12 | 2 | 0 | 0 | - |
| VP-5 | M | no | 1x | B.1.1.7 | 10 | 3 | 3 | 0 | - |
| VP-6 | M | no | 1x | B.1.1.7 | 9 | 3 | 3 | 1.62 | - |
| VP-10 | F | no | 1x | B.1.620 | 16 | 4 | 1 | 0.91 | - |
| VP-12 | F | no | 1x | B.1.1.7 | 104 | 12 | 9 | 0.42 | P9L, D405N |
| VP-13 | F | no | 1x | B.1.1.7 | 16 | 4 | 3 | 0 | - |
| VP-14 | F | no | 1x | B.1.1.7 | 21 | 4 | 6 | 0 | - |
| VP-15 | M | no | 1x | B.1.1.7 | 16 | 4 | 4 | 0 | - |
| VP-19 | F | yes | 1x | B.1.1.7 | 17 | 3 | 1 | 0.86 | - |

[1] CP, convalescent plasma (from COVID-19 recovered donors); VP, vaccinated plasma (from non-COVID-19 healthy donors after the second injection of a mRNA-based vaccine). Patients (post CP or VP) presenting an anti-S IgG decay are highlighted in grey. [2] Number of intra-host mutations supported by at least 10% of the reads that differ between sequenced genomes of the same patient at any timepoint. [3] The rate of mutations reaching fixation (>70% of the reads) between the first and last sequenced samples was compared to the theoretical SARS-CoV-2 mutation rate of approximately 25 mutations per year. [4] Over time development of non-silent spike mutations. Five mutations were present within the receptor-binding domain (spanning from the 319 to 529 amino acid sequence). The underlined mutations have been described as potentially related to immune escape/evasion.

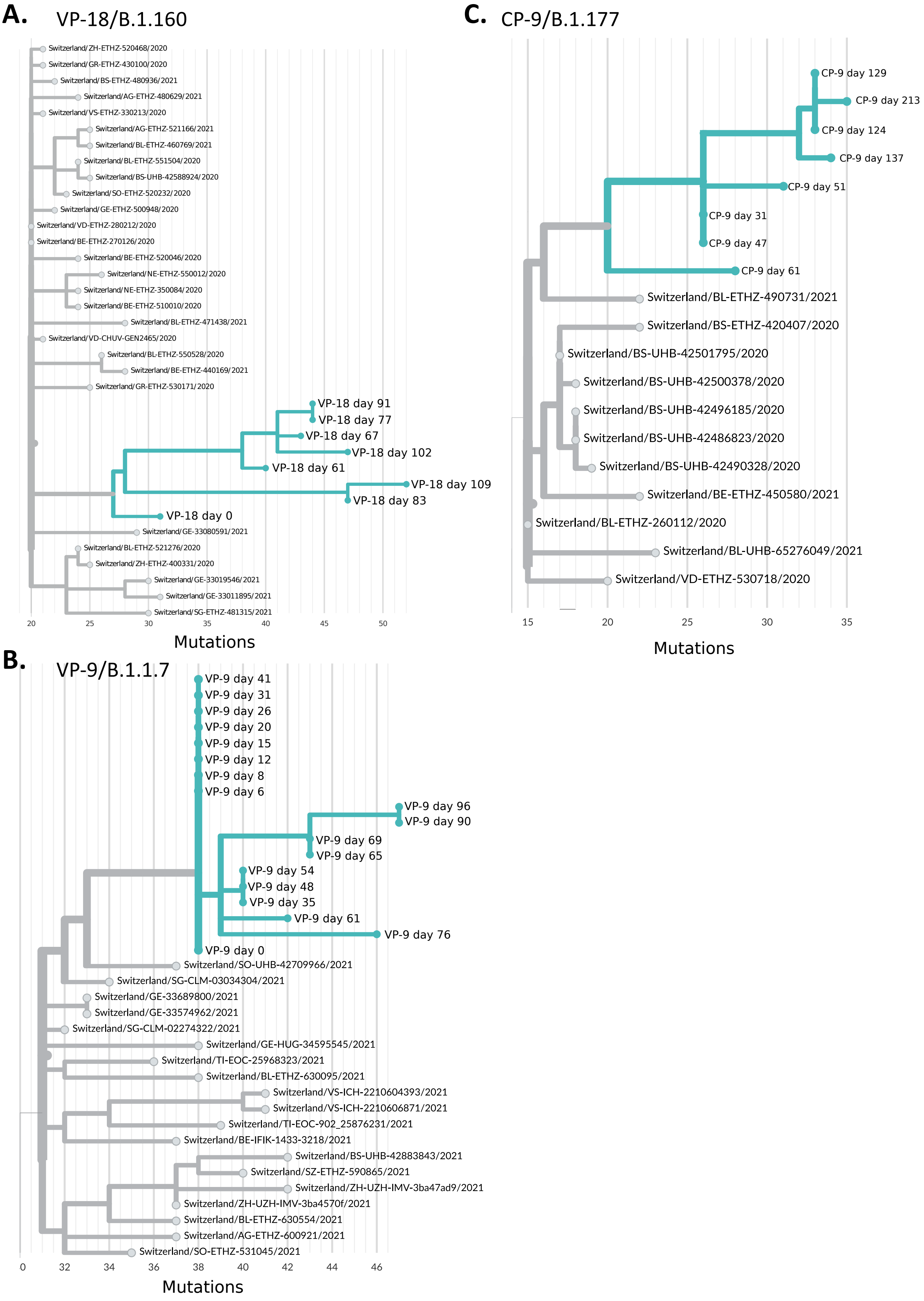


Supp Figure 1 Gachoud et al.

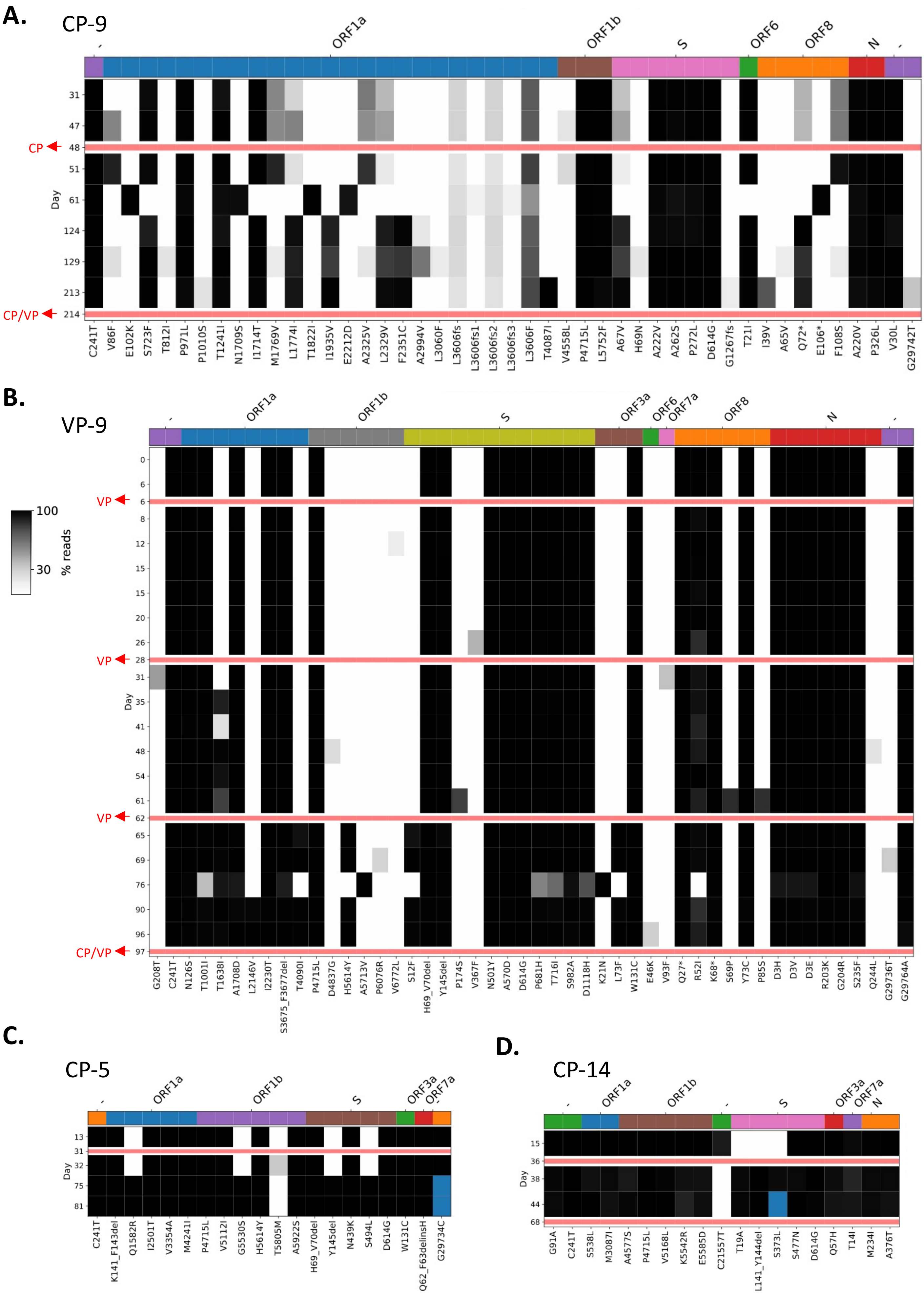


Supp Figure 2 Gachoud et al.

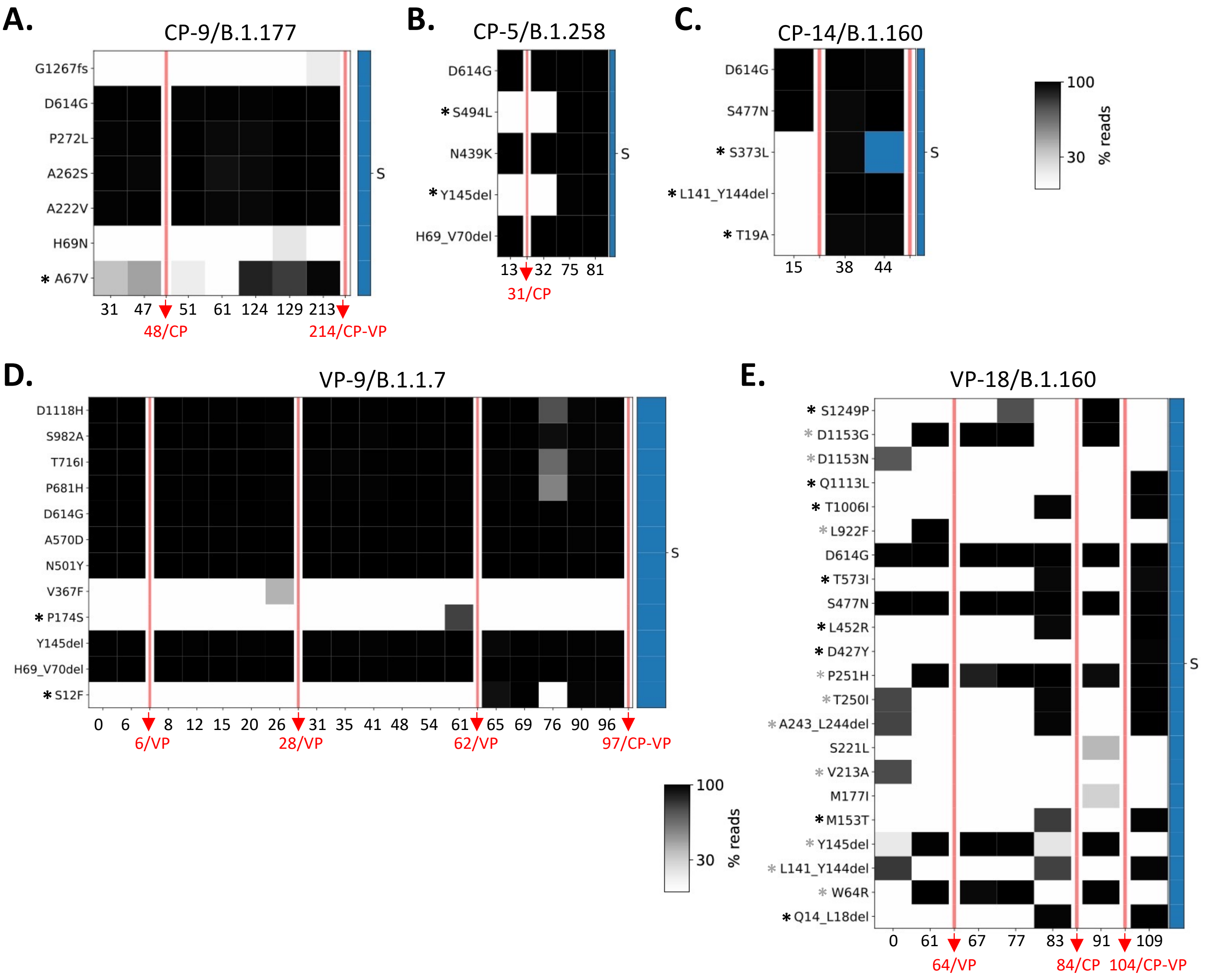


Supp Figure 3 Gachoud et al.

We gratefully acknowledge the following Authors from the Originating laboratories responsible for obtaining the specimens and the Submitting laboratories where genetic sequence data were generated and shared via the GISAID Initiative, on which this research is based.
All submitters of data may be contacted directly via www.gisaid.org
Shu, Y., McCauley, J. (2017) GISAID: from vision to reality EuroSurveillance 22(13) doi:10.2807/1560-7917.ES.2017.22.13.30494 PMCID: PMC5388101

| Virus name | Accession No. | Originating laboratory | Submitting laboratory | Authors |
|---|---|---|---|---|
| Switzerland/VS-ICH-2210604393/2021 | EPI_ISL_2521374 | Valais Hospital, Central Institute | Valais Hospital, Central Institute | Alexis Dumoulin, Lorenzo Cerutti, Henri Pegeot, Melyssa Elies, Deborah Penet, Keith Harshman, Ioannis Xenarios, Emmanouil Dermitzakis |
| Switzerland/VS-ICH-2210606871/2021 | EPI_ISL_2521377 | Valais Hospital, Central Institute | Valais Hospital, Central Institute | Alexis Dumoulin, Lorenzo Cerutti, Henri Pegeot, Melyssa Elies, Deborah Penet, Keith Harshman, Ioannis Xenarios, Emmanouil Dermitzakis |
| Switzerland/BL-ETHZ-521276/2020 | EPI_ISL_1260105 | Viollier AG | Department of Biosystems Science and Engineering, ETH Zürich | Chaoran Chen, Sarah Nadeau, Catharine Aquino, Ivan Topolsky, Philipp Jablonski, Lara Fuhrmann, David Dreifuss, Katharina Jahn, Andreia Cabral de Gouvea, Maria Domenica Moccia, Simon Grüter, Timothy Sykes, Lennart Opitz, Griffin White, Laura Neff, Doris Popovic, Andrea Patrignani, Jay Tracy, Ralph Schlapbach, Christiane Beckmann, Maurice Redondo, Olivier Kobel, Christoph Noppen, Sophie Seidel, Noemie Santamaria de Souza, Niko Beerenwinkel, Tanja Stadler |
| Switzerland/SG-ETHZ-481315/2021 | EPI_ISL_1059751 | Viollier AG | Department of Biosystems Science and Engineering, ETH Zürich | Chaoran Chen, Sarah Nadeau, Catharine Aquino, Ivan Topolsky, Philipp Jablonski, Lara Fuhrmann, David Dreifuss, Katharina Jahn, Andreia Cabral de Gouvea, Maria Domenica Moccia, Simon Grüter, Timothy Sykes, Lennart Opitz, Griffin White, Laura Neff, Doris Popovic, Andrea Patrignani, Jay Tracy, Ralph Schlapbach, Christiane Beckmann, Maurice Redondo, Olivier Kobel, Christoph Noppen, Sophie Seidel, Noemie Santamaria de Souza, Niko Beerenwinkel, Tanja Stadler |
| Switzerland/BL-ETHZ-490731/2021 | EPI_ISL_1059432 | Viollier AG | Department of Biosystems Science and Engineering, ETH Zürich | Chaoran Chen, Sarah Nadeau, Catharine Aquino, Ivan Topolsky, Philipp Jablonski, Lara Fuhrmann, David Dreifuss, Katharina Jahn, Andreia Cabral de Gouvea, Maria Domenica Moccia, Simon Grüter, Timothy Sykes, Lennart Opitz, Griffin White, Laura Neff, Doris Popovic, Andrea Patrignani, Jay Tracy, Ralph Schlapbach, Christiane Beckmann, Maurice Redondo, Olivier Kobel, Christoph Noppen, Sophie Seidel, Noemie Santamaria de Souza, Niko Beerenwinkel, Tanja Stadler |
| Switzerland/BS-ETHZ-420407/2020 | EPI_ISL_1260202 | Viollier AG | Department of Biosystems Science and Engineering, ETH Zürich | Chaoran Chen, Sarah Nadeau, Catharine Aquino, Ivan Topolsky, Philipp Jablonski, Lara Fuhrmann, David Dreifuss, Katharina Jahn, Andreia Cabral de Gouvea, Maria Domenica Moccia, Simon Grüter, Timothy Sykes, Lennart Opitz, Griffin White, Laura Neff, Doris Popovic, Andrea Patrignani, Jay Tracy, Ralph Schlapbach, Christiane Beckmann, Maurice Redondo, Olivier Kobel, Christoph Noppen, Sophie Seidel, Noemie Santamaria de Souza, Niko Beerenwinkel, Tanja Stadler |
| Switzerland/BE-ETHZ-450580/2021 | EPI_ISL_1002484 | Viollier AG | Department of Biosystems Science and Engineering, ETH Zürich | Chaoran Chen, Sarah Nadeau, Catharine Aquino, Ivan Topolsky, Philipp Jablonski, Lara Fuhrmann, David Dreifuss, Katharina Jahn, Andreia Cabral de Gouvea, Maria Domenica Moccia, Simon Grüter, Timothy Sykes, Lennart Opitz, Griffin White, Laura Neff, Doris Popovic, Andrea Patrignani, Jay Tracy, Ralph Schlapbach, Christiane Beckmann, Maurice Redondo, Olivier Kobel, Christoph Noppen, Sophie Seidel, Noemie Santamaria de Souza, Niko Beerenwinkel, Tanja Stadler |
| Switzerland/VD-ETHZ-530718/2020 | EPI_ISL_1361307 | Viollier AG | Department of Biosystems Science and Engineering, ETH Zürich | Chaoran Chen, Sarah Nadeau, Catharine Aquino, Ivan Topolsky, Philipp Jablonski, Lara Fuhrmann, David Dreifuss, Katharina Jahn, Andreia Cabral de Gouvea, Maria Domenica Moccia, Simon Grüter, Timothy Sykes, Lennart Opitz, Griffin White, Laura Neff, Doris Popovic, Andrea Patrignani, Jay Tracy, Ralph Schlapbach, Christiane Beckmann, Maurice Redondo, Olivier Kobel, Christoph Noppen, Sophie Seidel, Noemie Santamaria de Souza, Niko Beerenwinkel, Tanja Stadler |

| | | | | |
|---|---|---|---|---|
| Switzerland/SZ-ETHZ-590865/2021 | EPI_ISL_1913649 | Viollier AG | Department of Biosystems Science and Engineering, ETH Zurich | Chaoran Chen, Sarah Nadeau, Catharine Aquino, Ivan Topolsky, Philipp Jablonski, Lara Fuhrmann, David Dreifuss, Katharina Jahn, Andreia Cabral de Gouvea, Maria Domenica Moccia, Simon Grüter, Timothy Sykes, Lennart Opitz, Griffin White, Laura Neff, Doris Popovic, Andrea Patrignani, Jay Tracy, Ralph Schlapbach, Christiane Beckmann, Maurice Redondo, Olivier Kobel, Christoph Noppen, Sophie Seidel, Noemie Santamaria de Souza, Niko Beerenwinkel, Tanja Stadler |
| Switzerland/BL-ETHZ-630554/2021 | EPI_ISL_2375294 | Viollier AG | Department of Biosystems Science and Engineering, ETH Zürich | Chaoran Chen, Sarah Nadeau, Catharine Aquino, Ivan Topolsky, Philipp Jablonski, Lara Fuhrmann, David Dreifuss, Katharina Jahn, Andreia Cabral de Gouvea, Maria Domenica Moccia, Simon Grüter, Timothy Sykes, Lennart Opitz, Griffin White, Laura Neff, Doris Popovic, Andrea Patrignani, Jay Tracy, Ralph Schlapbach, Christiane Beckmann, Maurice Redondo, Olivier Kobel, Christoph Noppen, Sophie Seidel, Noemie Santamaria de Souza, Niko Beerenwinkel, Tanja Stadler |
| Switzerland/AG-ETHZ-600921/2021 | EPI_ISL_2019038 | Viollier AG | Department of Biosystems Science and Engineering, ETH Zürich | Chaoran Chen, Sarah Nadeau, Catharine Aquino, Ivan Topolsky, Philipp Jablonski, Lara Fuhrmann, David Dreifuss, Katharina Jahn, Andreia Cabral de Gouvea, Maria Domenica Moccia, Simon Grüter, Timothy Sykes, Lennart Opitz, Griffin White, Laura Neff, Doris Popovic, Andrea Patrignani, Jay Tracy, Ralph Schlapbach, Christiane Beckmann, Maurice Redondo, Olivier Kobel, Christoph Noppen, Sophie Seidel, Noemie Santamaria de Souza, Niko Beerenwinkel, Tanja Stadler |
| Switzerland/SO-ETHZ-531045/2021 | EPI_ISL_1361168 | Viollier AG | Department of Biosystems Science and Engineering, ETH Zürich | Chaoran Chen, Sarah Nadeau, Catharine Aquino, Ivan Topolsky, Philipp Jablonski, Lara Fuhrmann, David Dreifuss, Katharina Jahn, Andreia Cabral de Gouvea, Maria Domenica Moccia, Simon Grüter, Timothy Sykes, Lennart Opitz, Griffin White, Laura Neff, Doris Popovic, Andrea Patrignani, Jay Tracy, Ralph Schlapbach, Christiane Beckmann, Maurice Redondo, Olivier Kobel, Christoph Noppen, Sophie Seidel, Noemie Santamaria de Souza, Niko Beerenwinkel, Tanja Stadler |
| Switzerland/ZH-ETHZ-520468/2020 | EPI_ISL_1408838 | Viollier AG | Department of Biosystems Science and Engineering, ETH Zürich | Chaoran Chen, Sarah Nadeau, Catharine Aquino, Ivan Topolsky, Philipp Jablonski, Lara Fuhrmann, David Dreifuss, Katharina Jahn, Andreia Cabral de Gouvea, Maria Domenica Moccia, Simon Grüter, Timothy Sykes, Lennart Opitz, Griffin White, Laura Neff, Doris Popovic, Andrea Patrignani, Jay Tracy, Ralph Schlapbach, Christiane Beckmann, Maurice Redondo, Olivier Kobel, Christoph Noppen, Sophie Seidel, Noemie Santamaria de Souza, Niko Beerenwinkel, Tanja Stadler |
| Switzerland/GR-ETHZ-430100/2020 | EPI_ISL_796442 | Viollier AG | Department of Biosystems Science and Engineering, ETH Zürich | Chaoran Chen, Sarah Nadeau, Catharine Aquino, Ivan Topolsky, Philipp Jablonski, Lara Fuhrmann, David Dreifuss, Katharina Jahn, Andreia Cabral de Gouvea, Maria Domenica Moccia, Simon Grüter, Timothy Sykes, Lennart Opitz, Griffin White, Laura Neff, Doris Popovic, Andrea Patrignani, Jay Tracy, Ralph Schlapbach, Christiane Beckmann, Maurice Redondo, Olivier Kobel, Christoph Noppen, Sophie Seidel, Noemie Santamaria de Souza, Niko Beerenwinkel, Tanja Stadler |
| Switzerland/AG-ETHZ-480629/2021 | EPI_ISL_1004129 | Viollier AG | Department of Biosystems Science and Engineering, ETH Zürich | Chaoran Chen, Sarah Nadeau, Catharine Aquino, Ivan Topolsky, Philipp Jablonski, Lara Fuhrmann, David Dreifuss, Katharina Jahn, Andreia Cabral de Gouvea, Maria Domenica Moccia, Simon Grüter, Timothy Sykes, Lennart Opitz, Griffin White, Laura Neff, Doris Popovic, Andrea Patrignani, Jay Tracy, Ralph Schlapbach, Christiane Beckmann, Maurice Redondo, Olivier Kobel, Christoph Noppen, Sophie Seidel, Noemie Santamaria de Souza, Niko Beerenwinkel, Tanja Stadler |

| | | | | |
|---|---|---|---|---|
| Switzerland/AG-ETHZ-521166/2021 | EPI_ISL_1408495 | Viollier AG | Department of Biosystems Science and Engineering, ETH Zürich | Chaoran Chen, Sarah Nadeau, Catharine Aquino, Ivan Topolsky, Philipp Jablonski, Lara Fuhrmann, David Dreifuss, Katharina Jahn, Andreia Cabral de Gouvea, Maria Domenica Moccia, Simon Grüter, Timothy Sykes, Lennart Opitz, Griffin White, Laura Neff, Doris Popovic, Andrea Patrignani, Jay Tracy, Ralph Schlapbach, Christiane Beckmann, Maurice Redondo, Olivier Kobel, Christoph Noppen, Sophie Seidel, Noemie Santamaria de Souza, Niko Beerenwinkel, Tanja Stadler |
| Switzerland/BL-ETHZ-460769/2021 | EPI_ISL_1002987 | Viollier AG | Department of Biosystems Science and Engineering, ETH Zürich | Chaoran Chen, Sarah Nadeau, Catharine Aquino, Ivan Topolsky, Philipp Jablonski, Lara Fuhrmann, David Dreifuss, Katharina Jahn, Andreia Cabral de Gouvea, Maria Domenica Moccia, Simon Grüter, Timothy Sykes, Lennart Opitz, Griffin White, Laura Neff, Doris Popovic, Andrea Patrignani, Jay Tracy, Ralph Schlapbach, Christiane Beckmann, Maurice Redondo, Olivier Kobel, Christoph Noppen, Sophie Seidel, Noemie Santamaria de Souza, Niko Beerenwinkel, Tanja Stadler |
| Switzerland/BL-ETHZ-551504/2020 | EPI_ISL_1496031 | Viollier AG | Department of Biosystems Science and Engineering, ETH Zürich | Chaoran Chen, Sarah Nadeau, Catharine Aquino, Ivan Topolsky, Philipp Jablonski, Lara Fuhrmann, David Dreifuss, Katharina Jahn, Andreia Cabral de Gouvea, Maria Domenica Moccia, Simon Grüter, Timothy Sykes, Lennart Opitz, Griffin White, Laura Neff, Doris Popovic, Andrea Patrignani, Jay Tracy, Ralph Schlapbach, Christiane Beckmann, Maurice Redondo, Olivier Kobel, Christoph Noppen, Sophie Seidel, Noemie Santamaria de Souza, Niko Beerenwinkel, Tanja Stadler |
| Switzerland/GE-ETHZ-500948/2020 | EPI_ISL_1119495 | Viollier AG | Department of Biosystems Science and Engineering, ETH Zürich | Chaoran Chen, Sarah Nadeau, Catharine Aquino, Ivan Topolsky, Philipp Jablonski, Lara Fuhrmann, David Dreifuss, Katharina Jahn, Andreia Cabral de Gouvea, Maria Domenica Moccia, Simon Grüter, Timothy Sykes, Lennart Opitz, Griffin White, Laura Neff, Doris Popovic, Andrea Patrignani, Jay Tracy, Ralph Schlapbach, Christiane Beckmann, Maurice Redondo, Olivier Kobel, Christoph Noppen, Sophie Seidel, Noemie Santamaria de Souza, Niko Beerenwinkel, Tanja Stadler |
| Switzerland/BE-ETHZ-510010/2020 | EPI_ISL_1194895 | Viollier AG | Department of Biosystems Science and Engineering, ETH Zürich | Chaoran Chen, Sarah Nadeau, Catharine Aquino, Ivan Topolsky, Philipp Jablonski, Lara Fuhrmann, David Dreifuss, Katharina Jahn, Andreia Cabral de Gouvea, Maria Domenica Moccia, Simon Grüter, Timothy Sykes, Lennart Opitz, Griffin White, Laura Neff, Doris Popovic, Andrea Patrignani, Jay Tracy, Ralph Schlapbach, Christiane Beckmann, Maurice Redondo, Olivier Kobel, Christoph Noppen, Sophie Seidel, Noemie Santamaria de Souza, Niko Beerenwinkel, Tanja Stadler |
| Switzerland/BL-ETHZ-471438/2021 | EPI_ISL_1059532 | Viollier AG | Department of Biosystems Science and Engineering, ETH Zürich | Chaoran Chen, Sarah Nadeau, Catharine Aquino, Ivan Topolsky, Philipp Jablonski, Lara Fuhrmann, David Dreifuss, Katharina Jahn, Andreia Cabral de Gouvea, Maria Domenica Moccia, Simon Grüter, Timothy Sykes, Lennart Opitz, Griffin White, Laura Neff, Doris Popovic, Andrea Patrignani, Jay Tracy, Ralph Schlapbach, Christiane Beckmann, Maurice Redondo, Olivier Kobel, Christoph Noppen, Sophie Seidel, Noemie Santamaria de Souza, Niko Beerenwinkel, Tanja Stadler |
| Switzerland/BL-ETHZ-550528/2020 | EPI_ISL_1598100 | Viollier AG | Department of Biosystems Science and Engineering, ETH Zürich | Chaoran Chen, Sarah Nadeau, Catharine Aquino, Ivan Topolsky, Philipp Jablonski, Lara Fuhrmann, David Dreifuss, Katharina Jahn, Andreia Cabral de Gouvea, Maria Domenica Moccia, Simon Grüter, Timothy Sykes, Lennart Opitz, Griffin White, Laura Neff, Doris Popovic, Andrea Patrignani, Jay Tracy, Ralph Schlapbach, Christiane Beckmann, Maurice Redondo, Olivier Kobel, Christoph Noppen, Sophie Seidel, Noemie Santamaria de Souza, Niko Beerenwinkel, Tanja Stadler |
| Switzerland/BS-ETHZ-480936/2021 | EPI_ISL_1002422 | Viollier AG | Department of Biosystems Science and Engineering, ETH Zürich | Christian Beisel, Sarah Nadeau, Chaoran Chen, Ivan Topolsky, Philipp Jablonski, Lara Fuhrmann, David Dreifuss, Katharina Jahn, Rebecca Denes, Ina Nissen, Natascha Santacroce, Elodie Burcklen, Christiane Beckmann, Maurice Redondo, Olivier Kobel, Christoph Noppen, Sophie Seidel, Noemie Santamaria de Souza, Niko Beerenwinkel, Tanja Stadler |

| | | | | |
|---|---|---|---|---|
| Switzerland/VS-ETHZ-330213/2020 | EPI_ISL_1496655 | Viollier AG | Department of Biosystems Science and Engineering, ETH Zürich | Christian Beisel, Sarah Nadeau, Chaoran Chen, Ivan Topolsky, Philipp Jablonski, Lara Fuhrmann, David Dreifuss, Katharina Jahn, Rebecca Denes, Ina Nissen, Natascha Santacroce, Elodie Burcklen, Christiane Beckmann, Maurice Redondo, Olivier Kobel, Christoph Noppen, Sophie Seidel, Noemie Santamaria de Souza, Niko Beerenwinkel, Tanja Stadler |
| Switzerland/SO-ETHZ-520232/2020 | EPI_ISL_1260409 | Viollier AG | Department of Biosystems Science and Engineering, ETH Zürich | Christian Beisel, Sarah Nadeau, Chaoran Chen, Ivan Topolsky, Philipp Jablonski, Lara Fuhrmann, David Dreifuss, Katharina Jahn, Rebecca Denes, Ina Nissen, Natascha Santacroce, Elodie Burcklen, Christiane Beckmann, Maurice Redondo, Olivier Kobel, Christoph Noppen, Sophie Seidel, Noemie Santamaria de Souza, Niko Beerenwinkel, Tanja Stadler |
| Switzerland/VD-ETHZ-280212/2020 | EPI_ISL_560526 | Viollier AG | Department of Biosystems Science and Engineering, ETH Zürich | Christian Beisel, Sarah Nadeau, Chaoran Chen, Ivan Topolsky, Philipp Jablonski, Lara Fuhrmann, David Dreifuss, Katharina Jahn, Rebecca Denes, Ina Nissen, Natascha Santacroce, Elodie Burcklen, Christiane Beckmann, Maurice Redondo, Olivier Kobel, Christoph Noppen, Sophie Seidel, Noemie Santamaria de Souza, Niko Beerenwinkel, Tanja Stadler |
| Switzerland/BE-ETHZ-270126/2020 | EPI_ISL_541433 | Viollier AG | Department of Biosystems Science and Engineering, ETH Zürich | Christian Beisel, Sarah Nadeau, Chaoran Chen, Ivan Topolsky, Philipp Jablonski, Lara Fuhrmann, David Dreifuss, Katharina Jahn, Rebecca Denes, Ina Nissen, Natascha Santacroce, Elodie Burcklen, Christiane Beckmann, Maurice Redondo, Olivier Kobel, Christoph Noppen, Sophie Seidel, Noemie Santamaria de Souza, Niko Beerenwinkel, Tanja Stadler |
| Switzerland/BE-ETHZ-520046/2020 | EPI_ISL_1260047 | Viollier AG | Department of Biosystems Science and Engineering, ETH Zürich | Christian Beisel, Sarah Nadeau, Chaoran Chen, Ivan Topolsky, Philipp Jablonski, Lara Fuhrmann, David Dreifuss, Katharina Jahn, Rebecca Denes, Ina Nissen, Natascha Santacroce, Elodie Burcklen, Christiane Beckmann, Maurice Redondo, Olivier Kobel, Christoph Noppen, Sophie Seidel, Noemie Santamaria de Souza, Niko Beerenwinkel, Tanja Stadler |
| Switzerland/NE-ETHZ-550012/2020 | EPI_ISL_1598999 | Viollier AG | Department of Biosystems Science and Engineering, ETH Zürich | Christian Beisel, Sarah Nadeau, Chaoran Chen, Ivan Topolsky, Philipp Jablonski, Lara Fuhrmann, David Dreifuss, Katharina Jahn, Rebecca Denes, Ina Nissen, Natascha Santacroce, Elodie Burcklen, Christiane Beckmann, Maurice Redondo, Olivier Kobel, Christoph Noppen, Sophie Seidel, Noemie Santamaria de Souza, Niko Beerenwinkel, Tanja Stadler |
| Switzerland/NE-ETHZ-350084/2020 | EPI_ISL_1496561 | Viollier AG | Department of Biosystems Science and Engineering, ETH Zürich | Christian Beisel, Sarah Nadeau, Chaoran Chen, Ivan Topolsky, Philipp Jablonski, Lara Fuhrmann, David Dreifuss, Katharina Jahn, Rebecca Denes, Ina Nissen, Natascha Santacroce, Elodie Burcklen, Christiane Beckmann, Maurice Redondo, Olivier Kobel, Christoph Noppen, Sophie Seidel, Noemie Santamaria de Souza, Niko Beerenwinkel, Tanja Stadler |
| Switzerland/BE-ETHZ-440169/2021 | EPI_ISL_899318 | Viollier AG | Department of Biosystems Science and Engineering, ETH Zürich | Christian Beisel, Sarah Nadeau, Chaoran Chen, Ivan Topolsky, Philipp Jablonski, Lara Fuhrmann, David Dreifuss, Katharina Jahn, Rebecca Denes, Ina Nissen, Natascha Santacroce, Elodie Burcklen, Christiane Beckmann, Maurice Redondo, Olivier Kobel, Christoph Noppen, Sophie Seidel, Noemie Santamaria de Souza, Niko Beerenwinkel, Tanja Stadler |
| Switzerland/GR-ETHZ-530171/2020 | EPI_ISL_1360923 | Viollier AG | Department of Biosystems Science and Engineering, ETH Zürich | Christian Beisel, Sarah Nadeau, Chaoran Chen, Ivan Topolsky, Philipp Jablonski, Lara Fuhrmann, David Dreifuss, Katharina Jahn, Rebecca Denes, Ina Nissen, Natascha Santacroce, Elodie Burcklen, Christiane Beckmann, Maurice Redondo, Olivier Kobel, Christoph Noppen, Sophie Seidel, Noemie Santamaria de Souza, Niko Beerenwinkel, Tanja Stadler |
| Switzerland/ZH-ETHZ-400331/2020 | EPI_ISL_729165 | Viollier AG | Department of Biosystems Science and Engineering, ETH Zürich | Christian Beisel, Sarah Nadeau, Chaoran Chen, Ivan Topolsky, Philipp Jablonski, Lara Fuhrmann, David Dreifuss, Katharina Jahn, Rebecca Denes, Ina Nissen, Natascha Santacroce, Elodie Burcklen, Christiane Beckmann, Maurice Redondo, Olivier Kobel, Christoph Noppen, Sophie Seidel, Noemie Santamaria de Souza, Niko Beerenwinkel, Tanja Stadler |
| Switzerland/BL-ETHZ-260112/2020 | EPI_ISL_539421 | Viollier AG | Department of Biosystems Science and Engineering, ETH Zürich | Christian Beisel, Sarah Nadeau, Chaoran Chen, Ivan Topolsky, Philipp Jablonski, Lara Fuhrmann, David Dreifuss, Katharina Jahn, Rebecca Denes, Ina Nissen, Natascha Santacroce, Elodie Burcklen, Christiane Beckmann, Maurice Redondo, Olivier Kobel, Christoph Noppen, Sophie Seidel, Noemie Santamaria de Souza, Niko Beerenwinkel, Tanja Stadler |

| | | | | |
|---|---|---|---|---|
| Switzerland/BL-ETHZ-630095/2021 | EPI_ISL_2375116 | Viollier AG | Department of Biosystems Science and Engineering, ETH Zürich | Christian Beisel, Sarah Nadeau, Chaoran Chen, Ivan Topolsky, Philipp Jablonski, Lara Fuhrmann, David Dreifuss, Katharina Jahn, Rebecca Denes, Ina Nissen, Natascha Santacroce, Elodie Burcklen, Christiane Beckmann, Maurice Redondo, Olivier Kobel, Christoph Noppen, Sophie Seidel, Noemie Santamaria de Souza, Niko Beerenwinkel, Tanja Stadler |
| Switzerland/ZH-UZH-IMV-3ba47ad9/2021 | EPI_ISL_2086829 | UniversitŠtsSpital ZŸrich | Institute of Medical Virology | Daniel Ehrsam, Isabel StŸrmer, Catharine Aquino, Joel Wirz, Weihong Qi, Hubert Rehrauer, Verena Kufner, Gabriela Ziltener, Maryam Zaheri, Stefan Schmutz, Annette AudigŽ, Maria GrŸnberg, Kevin Steiner, Jon Huder, Cyril Shah, Riccarda Capaul, Guido Bloemberg, JŸrg Bšni, Michael Huber, Alexandra Trkola |
| Switzerland/BS-UHB-42486823/2020 | EPI_ISL_830954 | Clinical Virology | Clinical Bacteriology | Madlen Stange, Alfredo Mari, Tim Roloff, Helena MB Seth-Smith, Michael Schweitzer, Myrta Brunner, Karoline Leuzinger, Kirstine K. Soegaard, Alexander Gensch, Sarah Tschudin-Sutter, Simon Fuchs, Julia Bielicki, Hans Pargger, Martin Siegemund, Christian Nickel, Roland Bingisser, Michael Osthoff, Stefano Bassetti, Rita Schneider-Sliwa, Manuel Battegay, Hans Hirsch, Adrian Egli |
| Switzerland/TI-EOC-25968323/2021 | EPI_ISL_2708961 | Laboratorio di Microbiologia | Laboratorio di Microbiologia | Martinetti Lucchini Gladys, Valeria Spina |
| Switzerland/TI-EOC-902_25876231/2021 | EPI_ISL_2117439 | Laboratorio di Microbiologia | Laboratorio di Microbiologia | Martinetti Lucchini Gladys, Valeria Spina |
| Switzerland/GE-33080591/2021 | EPI_ISL_953879 | University Hospitals of Geneva, Laboratory c | HUG, Laboratory of Virology and the Health2030 Genome Center | Samuel Cordey, Ana Rita Goncalves, Laurent Kaiser, Lorenzo Cerutti, Henri Pegeot, Melyssa Elies, Deborah Penet, Keith Harshman, Ioannis Xenarios, Emmanouil Dermitzakis |
| Switzerland/GE-33019546/2021 | EPI_ISL_953723 | University Hospitals of Geneva, Laboratory c | HUG, Laboratory of Virology and the Health2030 Genome Center | Samuel Cordey, Ana Rita Goncalves, Laurent Kaiser, Lorenzo Cerutti, Henri Pegeot, Melyssa Elies, Deborah Penet, Keith Harshman, Ioannis Xenarios, Emmanouil Dermitzakis |
| Switzerland/GE-33011895/2021 | EPI_ISL_897606 | University Hospitals of Geneva, Laboratory c | HUG, Laboratory of Virology and the Health2030 Genome Center | Samuel Cordey, Ana Rita Goncalves, Laurent Kaiser, Lorenzo Cerutti, Henri Pegeot, Melyssa Elies, Deborah Penet, Keith Harshman, Ioannis Xenarios, Emmanouil Dermitzakis |
| Switzerland/GE-33689800/2021 | EPI_ISL_1533323 | University Hospitals of Geneva, Laboratory c | HUG, Laboratory of Virology and the Health2030 Genome Center | Samuel Cordey, Ana Rita Goncalves, Laurent Kaiser, Lorenzo Cerutti, Henri Pegeot, Melyssa Elies, Deborah Penet, Keith Harshman, Ioannis Xenarios, Emmanouil Dermitzakis |
| Switzerland/GE-33574962/2021 | EPI_ISL_1369644 | University Hospitals of Geneva, Laboratory c | HUG, Laboratory of Virology and the Health2030 Genome Center | Samuel Cordey, Ana Rita Goncalves, Laurent Kaiser, Lorenzo Cerutti, Henri Pegeot, Melyssa Elies, Deborah Penet, Keith Harshman, Ioannis Xenarios, Emmanouil Dermitzakis |
| Switzerland/GE-HUG-34595545/2021 | EPI_ISL_2885483 | University Hospitals of Geneva, Laboratory c | HUG, Laboratory of Virology and the Health2030 Genome Center | Samuel Cordey, Ana Rita Goncalves, Laurent Kaiser, Lorenzo Cerutti, Henri Pegeot, Melyssa Elies, Deborah Penet, Keith Harshman, Ioannis Xenarios, Emmanouil Dermitzakis |
| Switzerland/BE-IFIK-1433-3218/2021 | EPI_ISL_2608337 | Institute for Infectious Diseases | Institute for Infectious Diseases | Stefan Neuenschwander, Christian Baumann, Miguel A Terrazos Miani, Cora SŠgesser, Pascal Bittel, Peter Keller, Franziska Suter-Riniker, Stephen L Leib, Alban Ramette |
| Switzerland/BS-UHB-42588924/2020 | EPI_ISL_1747647 | Felix-Platter Spital | Clinical Bacteriology | Tim Roloff, Madlen Stange, Helena MB Seth-Smith, Alfredo Mari, Karoline Leuzinger, Julia Bielicki, Manuel Battegay, Hans Hirsch, Adrian Egli |
| Switzerland/BS-UHB-42501795/2020 | EPI_ISL_931099 | Clinical Virology | Clinical Bacteriology | Tim Roloff, Madlen Stange, Helena MB Seth-Smith, Alfredo Mari, Karoline Leuzinger, Julia Bielicki, Manuel Battegay, Hans Hirsch, Adrian Egli |
| Switzerland/BS-UHB-42500378/2020 | EPI_ISL_931060 | Clinical Virology | Clinical Bacteriology | Tim Roloff, Madlen Stange, Helena MB Seth-Smith, Alfredo Mari, Karoline Leuzinger, Julia Bielicki, Manuel Battegay, Hans Hirsch, Adrian Egli |
| Switzerland/BS-UHB-42496185/2020 | EPI_ISL_1388781 | University Hospital Basel, Clinical Virology | University Hospital Basel, Clinical Bacteriology | Tim Roloff, Madlen Stange, Helena MB Seth-Smith, Alfredo Mari, Karoline Leuzinger, Julia Bielicki, Manuel Battegay, Hans Hirsch, Adrian Egli |
| Switzerland/BS-UHB-42490328/2020 | EPI_ISL_930888 | Clinical Virology | Clinical Bacteriology | Tim Roloff, Madlen Stange, Helena MB Seth-Smith, Alfredo Mari, Karoline Leuzinger, Julia Bielicki, Manuel Battegay, Hans Hirsch, Adrian Egli |
| Switzerland/BL-UHB-65276049/2021 | EPI_ISL_1273460 | Clinical Virology | Clinical Bacteriology | Tim Roloff, Madlen Stange, Helena MB Seth-Smith, Alfredo Mari, Karoline Leuzinger, Julia Bielicki, Manuel Battegay, Hans Hirsch, Adrian Egli |
| Switzerland/SO-UHB-42709966/2021 | EPI_ISL_1747618 | Viollier AG | Clinical Bacteriology | Tim Roloff, Madlen Stange, Helena MB Seth-Smith, Alfredo Mari, Karoline Leuzinger, Julia Bielicki, Manuel Battegay, Hans Hirsch, Adrian Egli |
| Switzerland/BS-UHB-42883843/2021 | EPI_ISL_2610998 | Clinical Virology | Clinical Bacteriology | Tim Roloff, Madlen Stange, Helena MB Seth-Smith, Alfredo Mari, Karoline Leuzinger, Julia Bielicki, Manuel Battegay, Hans Hirsch, Adrian Egli |

| | | | | |
|---|---|---|---|---|
| Switzerland/ZH-UZH-IMV-3ba4570f/2021 | EPI_ISL_1663694 | Spital Limmattal | Institute of Medical Virology | Verena Kufner, Gabriela Ziltener, Maryam Zaheri, Stefan Schmutz, Annette Audigé, Maria Grünberg, Kevin Steiner, Jon Huder, Cyril Shah, Riccarda Capaul, Guido Bloemberg, Jürg Böni, Michael Huber, Alexandra Trkola |
| Switzerland/SG-CLM-03034304/2021 | EPI_ISL_1594310 | Center for Laboratory Medicine St. Gallen | Center for Laboratory Medicine St. Gallen | Yannick Gerth |
| Switzerland/SG-CLM-02274322/2021 | EPI_ISL_1594304 | Center for Laboratory Medicine St. Gallen | Center for Laboratory Medicine St. Gallen | Yannick Gerth |
